# Identifying US Counties with High Cumulative COVID-19 Burden and Their Characteristics

**DOI:** 10.1101/2020.12.02.20234989

**Authors:** Daniel Li, Sheila M. Gaynor, Corbin Quick, Jarvis T. Chen, Briana J.K. Stephenson, Brent A. Coull, Xihong Lin

## Abstract

Identifying areas with high COVID-19 burden and their characteristics can help improve vaccine distribution and uptake, reduce burdens on health care systems, and allow for better allocation of public health intervention resources. Synthesizing data from various government and nonprofit institutions of 3,142 United States (US) counties as of 12/21/2020, we studied county-level characteristics that are associated with cumulative case and death rates using regression analyses. Our results showed counties that are more rural, counties with more White/non-White segregation, and counties with higher percentages of people of color, in poverty, with no high school diploma, and with medical comorbidities such as diabetes and hypertension are associated with higher cumulative COVID-19 case and death rates. We identify the hardest hit counties in US using model-estimated case and death rates, which provide more reliable estimates of cumulative COVID-19 burdens than those using raw observed county-specific rates. Identification of counties with high disease burdens and understanding the characteristics of these counties can help inform policies to improve vaccine distribution, deployment and uptake, prevent overwhelming health care systems, and enhance testing access, personal protection equipment access, and other resource allocation efforts, all of which can help save more lives for vulnerable communities.

**Significance statement:** We found counties that are more rural, counties with more White/non-White segregation, and counties with higher percentages of people of color, in poverty, with no high school diploma, and with medical comorbidities such as diabetes and hypertension are associated with higher cumulative COVID-19 case and death rates. We also identified individual counties with high cumulative COVID-19 burden. Identification of counties with high disease burdens and understanding the characteristics of these counties can help inform policies to improve vaccine distribution, deployment and uptake, prevent overwhelming health care systems, and enhance testing access, personal protection equipment access, and other resource allocation efforts, all of which can help save more lives for vulnerable communities.

## Introduction

COVID-19 has had significant medical, social, and economic impacts on all communities in the United States (US), and efficient allocation of limited resources, mitigation of public health control measures, and prevention of overwhelming health care systems have been constant challenges (1–8). Concerns about access to testing and personal protective equipment (PPE) arose during Spring 2020, and existing challenges in Winter 2020-2021 include vaccine distribution, deployment and uptake (9–14). The rapidly increasing numbers of COVID-19 cases, hospitalizations, and deaths in the US present an urgent need to develop efficient vaccine distribution, deployment and uptake strategies and to craft effective public health measures. This will allow for better control of the pandemic and reduce the burden of COVID-19 on health care systems.

Current US national vaccine guidelines use individual-level factors to prioritize certain high-risk people, such as healthcare workers and long-term care residents first, and then certain essential workers and elderly (15, 16). However, this strategy has been shown to lead to vaccines being distributed proportional to population density, leaving more sparsely populated areas at increased risk (17). Given the high vaccine efficacy of the Moderna and Pfizer-BioNTech COVID-19 vaccines, vaccination implementation and compliance to increase vaccine uptake is the defining challenge in 2021(18, 19). Vaccine distribution and uptake strategies that additionally account for spatial information and local community characteristics of regions with the most COVID-19 cases and deaths can help more rapidly get vaccines into people’s arms, save even more lives, and make the COVID-19 burden on healthcare systems more manageable (20–22).

It is hence of substantial interest to identify counties of high case and death rates and understand the characteristics of these counties. For a comprehensive assessment of total COVID-19 impact across all US regions, raw observed case and death counts and rates per capita (population size) at the county level are typically presented (23–25). However, 2019 US county population sizes vary from as small as 86 residents in Kalawao County, Hawaii to over 10 million residents in Los Angeles County, California. Using per capita rates, one case equivalent in Kalawao County is equivalent to approximately 117,000 cases in Los Angeles County. Crude rate estimates for small counties are unstable, and a more statistically sound framework is needed to appropriately quantify total COVID-19 case and death burdens for US counties of all population sizes. By leveraging data across different counties and accounting for uncertainty, and we can also identify county-level characteristics that are associated with county-level disease burdens.

In this study, we use county-level demographic, race/ethnicity, socioeconomic, and medical comorbidities variables synthesized from various data sources to build regression models that i) study county-level covariate associations with COVID-19 total case and death rates, ii) provide estimates of COVID-19 total case and death rates that are more reliable than raw observed rates for all US counties, and iii) explore season-varying county-level associations with COVID-19 weekly case and death rates. Our models show disease burdens greatly vary between counties, and our models reveal common characteristics of counties with high disease burdens. Counties that are more rural, have a greater percentage of people of color, more White/non-White segregation, and more residents in poverty, with no high school diploma, and/or with medical comorbidities, such as diabetes and hypertension, are associated with higher cumulative COVID-19 case and death rates.

Our models provide more reliable model-based county cumulative case and death rates by leveraging information across counties and accounting for instability in estimated rates from less populated counties. Season-varying analyses revealed that urban counties with more racial minorities were associated with higher weekly case and death rates in the Spring, more rural counties were associated with higher weekly rates in the Fall, and counties with more residents with no high school diploma and comorbidities such as hypertension were associated with higher weekly rates in the Spring, Summer, and Fall. These association analysis results and county-specific case and death rate estimates can be used to identify vulnerable US counties with high-risk county-level factors and with the greatest total COVID-19 burden to date. Such counties can be prioritized for prevention and intervention efforts including but not limited to vaccine distribution and uptake, testing and resources for control measures, and management of COVID-19 burdens on local health care systems.

### Brief Materials and Methods

We obtained demographic, socioeconomic and comorbidity data from a COVID-19 GitHub repository that drew from the US Department of Agriculture, Area Health Resources Files, County Health Rankings and Roadmaps, Centers for Disease Control and Prevention, and Kaiser News Health (26). We obtained COVID-19 county cases and deaths from 1/22/20-12/21/20 from USA Facts (27) and additional demographic data from the US Census Bureau (28).

We used a Poisson mixed model to model cumulative case and death counts for all 3,142 US counties through 12/21/20. We included fixed effects for each state to account for state-to-state variation not explained by variables in the model (such as state testing rates), a random effect for each county to account for overdispersion and county testing rates, and an offset term for log county population (2019 US Census estimates). Model variables are listed in **Table S1** and model covariate geographic heatmaps are in **Figures S1-S2**. Continuous covariates were scaled to have mean zero and standard deviation one for modeling. County model-based estimates and confidence intervals were calculated from model output.

Model coefficient estimates and confidence intervals were used to explore associations between county-level covariates and COVID-19 rates. Exploratory total case fatality rate (CFR) analyses were performed by modeling total county death counts with an offset for log total case counts. Additional spatiotemporal modeling of weekly county case, death, and case fatality rates from 3/23/20-12/21/20 (40 repeated measures per county) was performed to investigate how county-level associations varied by season. Extra details about data sources and statistical methods are available in the full Materials and Methods.

## Results

Our model estimated case and death rates matched well with the corresponding observed rates. **Figure1** shows heatmaps of the observed rates, heatmaps of the estimated rates, and a scatterplot of observed versus estimated values by county. There was the most agreement between observed and estimated case rates (**Figure 1A)**. While there were only 3 counties reporting zero total cases as of 12/21/20, there were 111 counties reporting zero total deaths as of 12/21/20. These zero death counties can be appreciated in the left panel observed heatmaps of **Figure 1B** by the “coarse, pixelated” patterns, particularly noticeable in the central Midwest. The corresponding center panel estimated death rate map values are noticeably “smoother” in this region. These patterns are also reflected in the right panel scatterplot by the increased number of points on the y-axis.

**Fig. 1.**
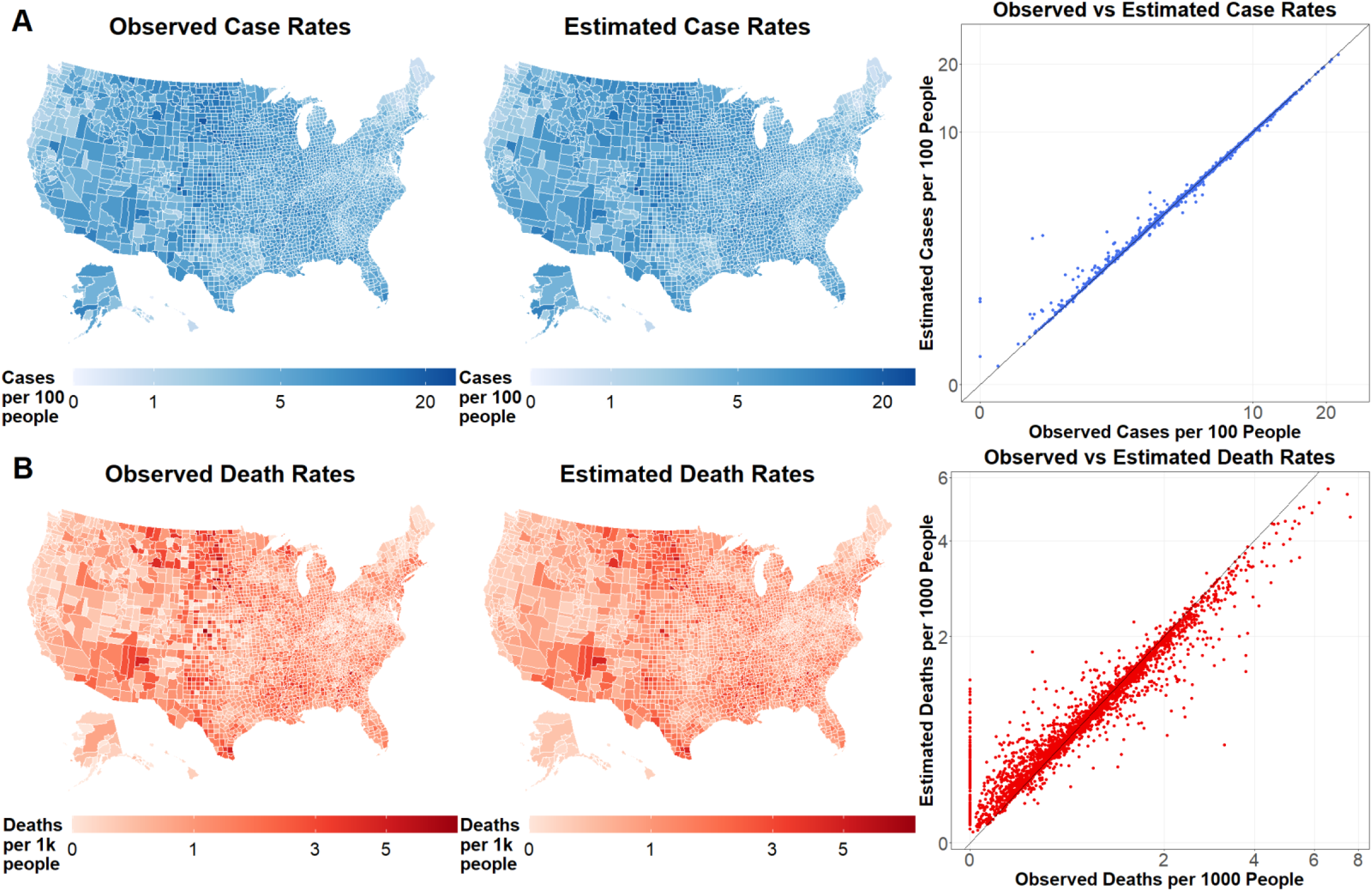
Observed and estimated case, death, and case fatality rates. Observed and multivariable model estimated cumulative (A) case rates and (B) death rates through 12/21/2020 for all 3,142 US counties. Left panels are heatmaps of observed rates. Middle panels are heatmaps of estimated rates. Right panels are scatterplots of observed vs estimated rates. The solid line (intercept zero slope one) indicates perfect agreement between observed and estimated rates. All rates are generally well estimated, though 3 counties reported 0 total cases and 111 counties reported 0 total deaths.

Associational relative risks (RR) for the multiplicative increase in county COVID-19 cumulative case, death, and case fatality rates were calculated for a one standard deviation increase of a county-level variable. **Figure 2** shows univariable and multivariable case rate RR, and **Figure 3** shows univariable and multivariable death rate RR. Univariable analyses show that more rural counties and counties with more racial minorities, socioeconomic disadvantage, and increased health comorbidities tend to have higher total case and death rates. After adjustment in multivariable analyses, more rural counties, and counties with increased White/non-White segregation, poverty, no high school diplomas, diabetes, heart failure, and hypertension tended to have higher total case and/or death rates.

**Fig. 2.**
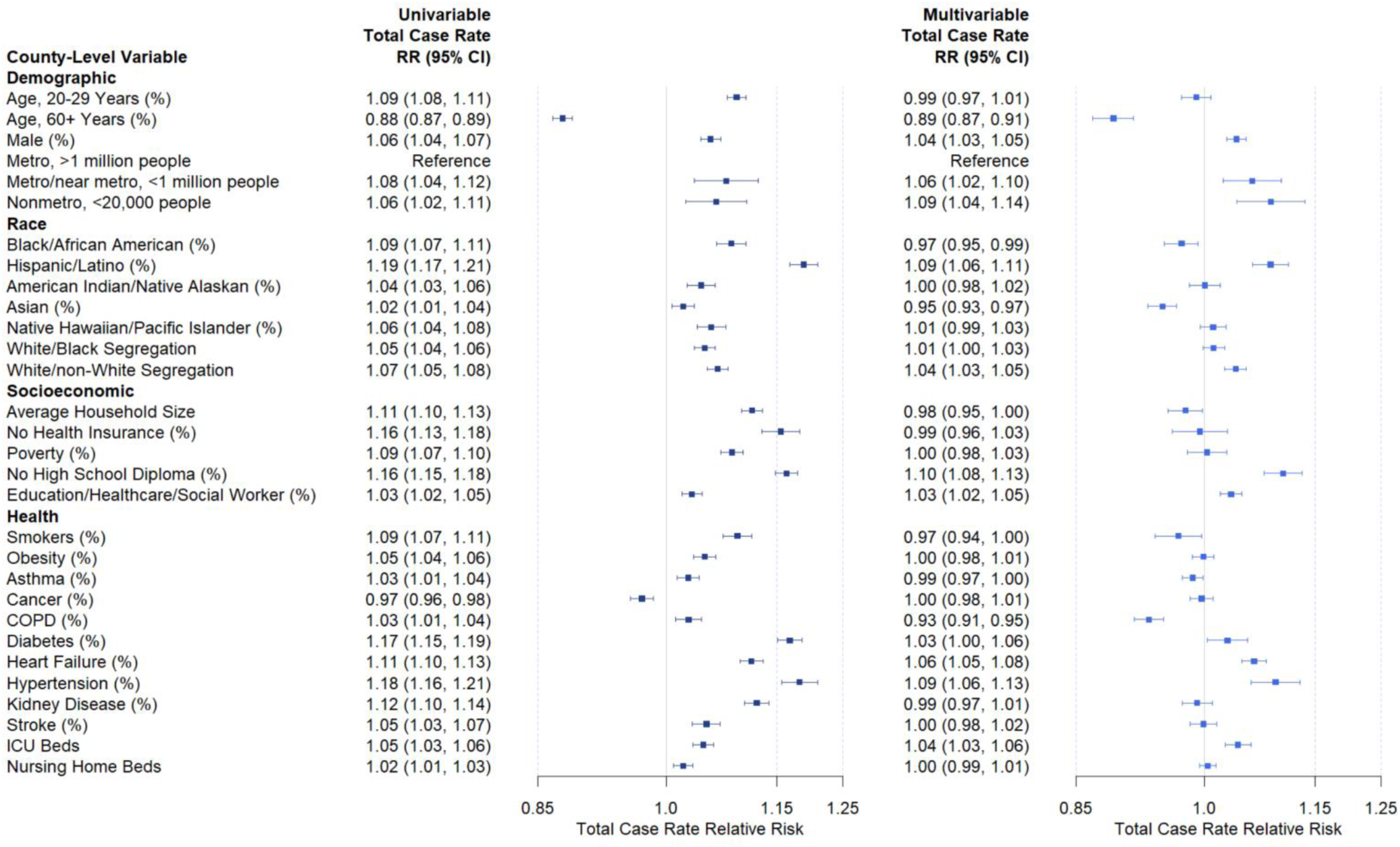
Univariable and multivariable case rate relative risks. Univariable and multivariable relative risks of demographic, socioeconomic, and health comorbidity factors on cumulative COVID-19 case rates through 12/21/20 additionally adjust for state fixed effects and county random effects. Boxes are point estimates and error bars mark 95% confidence intervals. Relative risks are for a one standard deviation increase in a variable (see **Table S1**), except for the metro/nonmetro categorical variable.

**Fig. 3.**
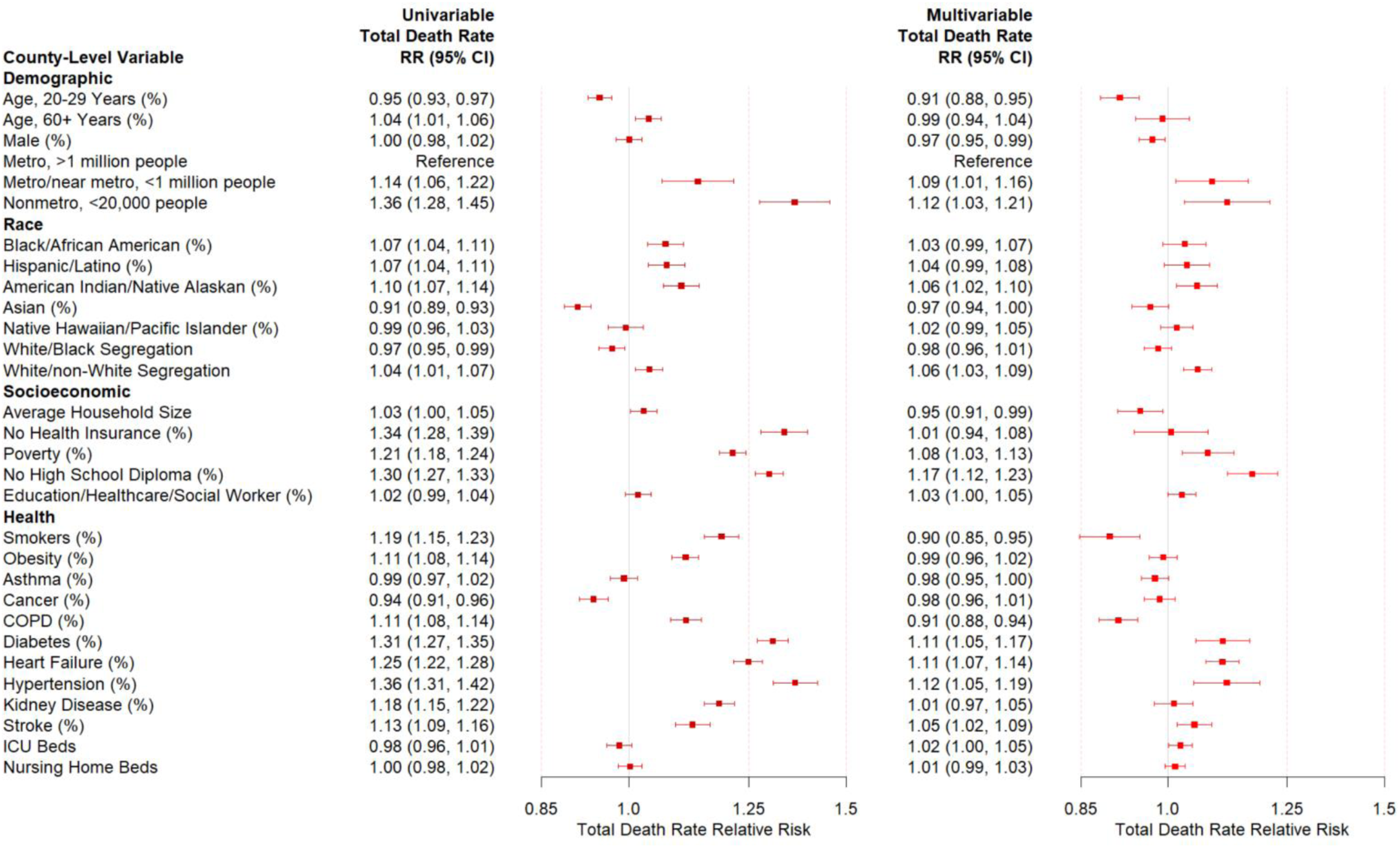
Univariable and multivariable death rate relative risks. Univariable and multivariable relative risks of demographic, socioeconomic, and health comorbidity factors on cumulative COVID-19 death rates through 12/21/20 additionally adjust for state fixed effects and county random effects. Boxes are point estimates and error bars mark 95% confidence intervals. Relative risks are for a one standard deviation increase in a variable (see **Table S1**), except for the metro/nonmetro categorical variable.

Many variables that were statistically significant on univariable analysis were not on multivariable analysis. **Figure S3** plots Spearman correlations for all covariates and shows how many of these variables are correlated. More rural counties tend to have more males residents, residents 60+ years, and fewer minorities. Black/African American percentage tended to be positively correlated with socioeconomic and health comorbidity variables. Socioeconomic and health comorbidity variables tended to be positively correlated. Spatial patterns among different covariates also exist. **Figures S4-S5** show heatmaps of standardized covariate values for modeling. Black percentages, stroke, heart failure, hypertension, and kidney disease percentages are all greatest in the Southern states. Hispanic percentages are greatest along the Southwestern states. Smoking and chronic obstructive pulmonary disease (COPD) percentages are elevated along the Appalachian Mountains.

Our model estimated rates can be used to identify states with the greatest COVID-19 burden. **Figure 4** shows (**A**) the counties with the top 30 estimated total case rates and (**B**) the counties with the top 30 estimated total death rates. States such as North Dakota, South Dakota, and Texas have multiple counties in the top 30 estimated case and/or death rates. Counties with smaller populations tend to have larger confidence intervals. For example, among case rates, Buffalo County in South Dakota has a population around 2,000 and a relatively larger confidence interval, while Hale County in Texas has a population around 33,000 and a relatively smaller confidence interval. Among death rates, Kenedy County in Texas has a population around 400 and a relatively larger confidence interval, while the Bronx Borough in New York has a population around 1.4 million and has a relatively smaller confidence interval.

**Fig. 4.**
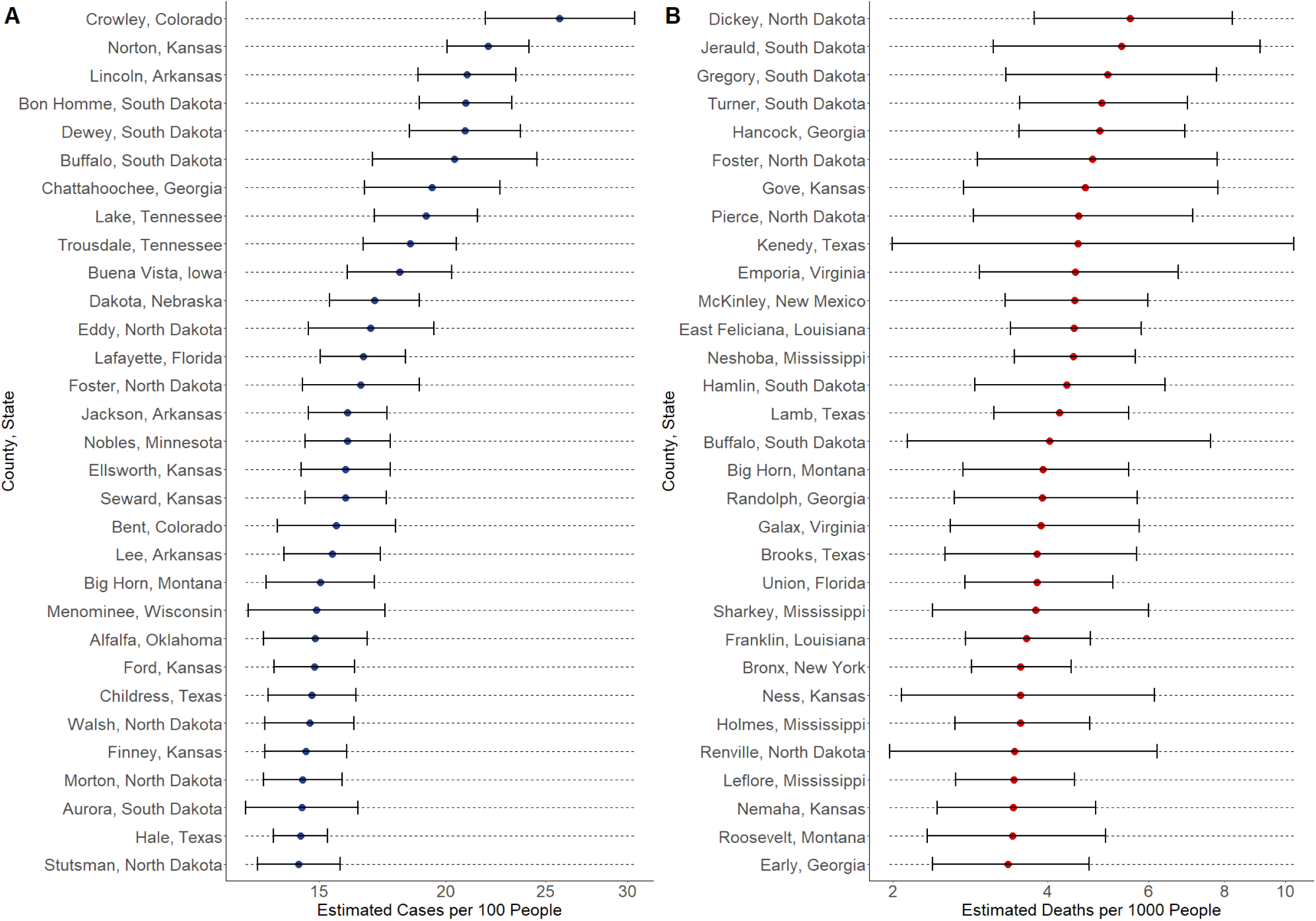
Identifying counties with greatest COVID-19 burden. Top 30 counties in the US with (A) the highest estimated case rates and (B) the highest estimated death rates. Circles are point estimates. Error bars are 95% model-based CIs.

We performed exploratory total case fatality rates (CFR) analyses. Assuming the ascertainment rates of reported cases vary by state or county, and by including fixed state effects and county random effects, the CFR and infection fatality rate (IFR) regression analyses produced identical results for covariate associations (see **Supporting Information** for further discussion). Observed and estimated CFR rates matched well, similar to the death rates (**Figure S6**). The CFR results had similar directions to the primary death rate results, and counties with more residents age 60+ years were associated with increased case fatality rates on both univariable and multivariable analyses (**Figure S7**). However, these analyses are likely subject to bias due to several factors, such as differential underestimation of the total number of cases by race and ethnicity selection bias of subjects who have been tested (29, 30).

We further explored how county-level covariate associations with COVID-19 rates varied over the course of the pandemic. **Figures S8-S9** show longitudinal trends of COVID-19 county cases and deaths over time. **Figures 8A** and **9A** show total case and death counts were mostly concentrated in a few areas including the Northeast during the Spring, increased in the South during the Summer, and increased everywhere including rural areas during the Fall. **Figures 8B** and **9B** show similar patterns for case and death rates respectively, though the increased case and death rates in some Midwest States are more prominent. **Figures 8C** and **9C** show case and death rates by US Census region were highest in the Northeast during the spring, highest in the South during the summer, and highest in the Midwest during the Fall.

Longitudinal modeling of weekly county case and death rates was performed. **Tables S2** and **S4** show univariable season varying relative risks for weekly case and death rates respectively, and **Tables S3** and **S5** show multivariable relative risks for weekly case and death rates respectively. Similar to total rate analyses, many covariates had significant associations in univariable modeling that changed in multivariable modeling. During the Spring, more urban counties and counties with more racial minorities and racial residential segregation tended to have greater case and death rates. As the pandemic progressed, during the Fall, more rural areas tended to have greater case and death rates. Socioeconomic variables such as no high school diploma and medical comorbidity variables such as hypertension were associated with increased case and death rates throughout the Fall, Summer, and Spring.

Lastly, we look at case fatality rates over time. **Figure S10** shows longitudinal trends of COVID-19 case fatality rates over time. **Figure S10A** shows how due to possible delays between COVID-19 testing confirmation and official recording of COVID-19 deaths, national death rates lagged by about one week peak at the same time as national case rates for USAFacts data. **Figure S10B** does not show any distinct geographic case fatality rate patterns, but case fatality rates appear to be decreasing in all regions over time. **Figures S10C** shows case fatality rates have fallen since the start of the pandemic.

Longitudinal modeling of case rates adjusted for the observed one-week reporting lag specific to USAFacts data (ex. weekly deaths ending on 12/21/20 were matched with weekly cases ending on 12/14/20). **Table S6** and **S7** show univariable and multivariable season-varying weekly case fatality relative risks. Counties with more residents ages 60+ years and counties with larger percentages of medical comorbidities tend to have increased case fatality relative risks throughout the Spring, Summer, and Fall. More rural areas have tended to have greater case fatality rates during the Fall. However, just as with the cumulative case fatality rate analyses, various reporting biases exist. Additional challenges include matching individuals death and case dates using aggregated data and instability from low weekly case count offsets, especially for sparsely populated areas.

## Discussion

Using county-level data obtained from various government and nonprofit sources, we provide model-based estimates of COVID-19 total case and death rates for every county in the US. Unlike observed per capita rates, our model-estimated rates are more stable in counties with smaller populations and allow for better quantification of estimate uncertainty. We also explore associations between county-level covariates and total case and death rates using our model, and we find that rural counties, counties with more White/non-White segregation, and counties with increased socioeconomic disadvantage and increased medical comorbidities all tend to have greater cumulative case and death rates. These findings and results can better inform policies for strategic vaccine distribution, deployment and uptake, mitigation of public health control measures, assessment of preparedness of health care system capacity, and reduction of health care burdens to vulnerable US counties.

The CDC’s Advisory Committee on Immunization Practices (ACIP) releases recommendations for vaccine distribution,(15, 16) but state governments ultimately decide how to distribute vaccines and encourage vaccine uptake (31). To assist states in formulating their own vaccine distribution policies and vaccine uptake strategies, we provide observed and estimated case, death, and case fatality rates for all US counties. Tailored strategies can also target county-level factors we have identified in associational analyses. All results will be updated regularly.

We are primarily interested in studying the associations between county-level covariates and total case and death rates to identify county-level characteristics for high disease burdens. These results can then be used to identify vulnerable counties so that tailored strategies can be developed to improve vaccine distribution and uptake and improve disease control measures. Our county-level association analysis results are consistent with results from various individual-level studies. Racial minorities have been found to have increased COVID-19 case, hospitalization, and death rates in the Spring (32–36). Older patients are more likely to develop severe COVID-19 symptoms and have greater mortality rates (37). Household size is known to affect COVID-19 contact and transmission rates (38). Heart failure, hypertension, and stroke are important biological and clinical risk factors for COVID-19 disease severity and mortality (37, 39).

We are primarily interested in this paper studying how county-level characteristics are associated with their cumulative case and death rates. These county-level associations should not be interpreted as individual level associations. Associations observed at an aggregated level may be in the same direction, different direction, or not exist at the individual level (40). As seen in **Figures S1-S2, S4-S5** and noted earlier in the results section, most county-level covariates have a clear geographic pattern that likely makes estimating individual-level effects difficult. Multiple covariates are correlated in multivariable modeling and complicate confounding issues in estimation and inference. Lastly, as with all observational studies, associational findings do not imply causality. However, the focus of our study is on estimating county-level case and death rates and identifying county-level characteristics of vulnerable counties to help counties develop tailored strategies for vaccination, public health control measure and management of health care burden, not estimating individual-level effects. Therefore, these limitations are less relevant to our primary study goals.

Our analyses of weekly COVID-19 case and death rates also support how COVID-19 has disproportionately affected various vulnerable populations at different times. The early stages of the pandemic in the Spring had disproportionately affected racially diverse but socioeconomically disadvantaged urban areas. More recently in the Fall, predominately white, rural, socioeconomically disadvantaged areas have been hit harder. Most research on the COVID-19 pandemic has focused on urban areas, and more studies of rural areas with individual-level data are needed to better characterize the experience of these diverse 46 million individuals (5, 41).

Next steps to improving our model-based estimates can include obtaining data at finer geographic resolution such as at the US census tract or zip code level. Additional variables such as neighborhood testing rates, percentage of healthcare workers, percentage of essential workers, exposure to infected individuals within households and in communities, personal protective equipment access, use of public transportation, utilization rates of available healthcare facilities/resources, access to living resources (such as a lack of access to clean water in many households of American Indian/Alaskan Native communities), health literacy, occupation and work conditions, housing conditions, basic living resources, and access to COVID-19 treatments may further improve modeling accuracy. As more counties report COVID-19 deaths and fewer counties have zero deaths, as with the case rates, our estimated death rates will likely also closer match the observed death rates. Modeling of longitudinal case and death counts can also be done using differential equations and incorporating information on unascertained and asymptomatic cases (29).

In summary, multi-faceted efforts are needed to combat the pandemic, optimize COVID-19 vaccine distribution, deployment and uptake and resource allocation, prevent the overload health care systems, and improve mitigation of control measures. Our model-estimated case and death rates, shared characteristics of counties with high disease rates, and identified spatiotemporal patterns of disease burdens can have a significant impact in making these decisions. Increased resources, such as vaccination education, testing priority and accessible points of care, should also be allocated to more rural counties and counties with more residential racial segregation, less education infrastructure, and/or greater prevalences of medical comorbidities. Intervention measures can include policies requiring face coverings, guaranteeing workers can take paid sick leave, providing personal protective equipment to essential workers, and ensuring prioritized and robust testing, tracing, and isolation infrastructure. Outreach efforts can include vaccination education by engaging community leaders and health care providers, transportation assistance, social and community support, and increased accessibility and affordability of health care. Our model estimated county case and death rates are more reliable metrics of total COVID-19 burden and can further help optimize resource allocation, plan education efforts on public control measure and vaccination deployment and uptake, reduce burdens on health care systems, and minimize additional loss of life.

## Materials and Methods

### Data Sources

USA Facts is a non-profit organization providing data about government tax revenues, expenditures, and outcomes.(27) Area Health Resources Files is a part of the federal government’s Health Resources & Services administrations that includes data on population characteristics, economics, hospital utilization, and more.(42) County Health Rankings & Roadmaps is a collaboration between the Robert Wood Johnson Foundation and University of Wisconsin that provides local community health data.(43)

### Outcome Variables

County-level cumulative and weekly COVID-19 cases and deaths as of 12/21/20 were directly obtained from USA Facts (44). USA Facts aggregates data from the Centers of Disease Control and state and local public health agencies. County-level data were confirmed by referencing state and local agencies.

### Covariates

Demographic variables were obtained from the US Census Bureau and US Department of Agriculture and included county percent ages 20-29 years, percent ages 60+ years, percent male, and metro/nonmetro status classification. US Department of Agriculture rural-urban continuum codes were grouped into three categories for the metro/nonmetro categorical variable: metro, population ≥ 1 million (code 1); metro or near metro, population 20,000 to 1 million (codes 2-4); nonmetro, population < 20,000 (codes 5-9) (45).

County-level population distribution by race/ethnicity, including Black/African American, Hispanic/Latino, American Indian/Native Alaskan, Asian, Native Hawaiian/Pacific Islander proportions, were directly obtained from 2019 US Census Bureau estimates (28). County residential racial segregation indices of dissimilarity were obtained from County Health Rankings & Roadmaps (46). These indices were originally calculated from data from US Census tracts from the American Community Survey 2014-2018. Counties with less than 100 Black/non-White residents had the index of dissimilarity set to be equal to 1.

Socioeconomic variables were obtained from Area Health Resource Files and included average household size, percentage of individuals between 18-64 years old without health insurance, percentage in poverty, percentage of people aged >25 years without a high school diploma, and percentage of people working in education/health care/social assistance.

Prevalence rates for several comorbidities were obtained from County Health Rankings & Roadmap. Comorbidities included county-level percentages for: smoking, obesity, asthma, cancer, chronic obstructive pulmonary disease, diabetes, heart failure, hypertension, kidney disease, and stroke. Kaiser News provided total intensive care unit (ICU) beds and nursing home beds.

Log transformations were applied to heavily skewed variables. Covariate information is summarized in **Table S1**, covariate heatmaps are in **Figures S1-S2**, and covariate Spearman correlations are presented in **Figures S3**.

### Statistical Analyses

Cumulative COVID-19 county case and county death counts were each modeled using a Poisson mixed effects model using log(county population size) as an offset. Models included fixed effects for covariates and indicator variables for each state to account for differences in state geography, testing rates, and other sources of variability. County specific random effects were included to account for overdispersion and county testing variability. All continuous variables were scaled to have mean 0 and standard deviation 1. Heatmaps of standardized covariates are in **Figures S4-S5**.

To explicitly define the cumulative case rate model, assume *Rij* is the number of reported cases and *P*_*ij*_ is the population of county *j* of state *i*. Then the case rate Poisson mixed model can be written as

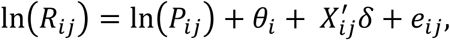

where *θ*_*i*_ is the state effect, *X*_*ij*_ is a vector of covariates, and the *e*_*ij*_ are county-specific random effects. The death and case fatality rate models follow the same structure. Total death rates modeled total deaths instead of total cases. Total case rates additionally used log total cases as an offset instead of log population size. For case fatality rate models, counties with total case counts of zero were set to one so the log offset would be defined.

Univariable models included a single covariate for *X*_*ij*_, and separate models were fit for each outcome and each covariate combination. Multivariable models included all covariates for *X*_*ij*_ and a single model was fit for each outcome. Predicted rates were calculated from the cumulative case and death rate models used. Predictions included estimated fixed and random effects (Best Linear Unbiased Prediction) and confidence intervals incorporated estimated standard errors for both fixed and random effects. Estimates and confidence intervals for *δ* were used to explore associations between county-level covariates and COVID-19 rates.

We also explored including state testing rates as a covariate for modeling. However, we found this did not change any estimated effects adjusted for demographic, race/ethnicity, socioeconomic, and comorbidity variables because we already controlled for fixed state effects using state dummy variables. Adding state testing rates only changed the estimated state fixed effects through re-parametrization.

To explore how county-level associations have changed with time, we modeled county weekly case and death counts (40 repeated measures per county). To define our case rate model, let *P*_*ij*_ be the population size and *R*_*ijt*_ be the observed deaths in county *j* of state *i* on week *t*. The Poisson mixed model with mean cases *λ*_*ijt*_ can be written as

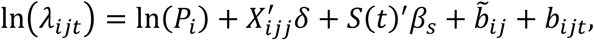

where *X*_*ijt*_ is a vector of covariates including state fixed effects and covariate season interactions, *δ* is a vector of regression coefficients, *S*(*t*)^′^*β*_*s*_ is a cubic spline basis for time with knots every 14 weeks that varies by US Census region, 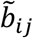 are independent and identically distributed (i.i.d.) county specific random effects with 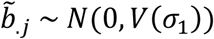, and *b*_*ijt*_ are county specific longitudinal random effects with AR-1 correlation structure *b*_*ij*_ i.i.d *N*(0, *V*(*σ*_2_)). Estimation and inference proceed using penalized quasi-likelihood as implemented in the “glmmpql” command from the MASS package in R.(47–49)

The weekly death rate model is the same except weekly new deaths instead of weekly case counts are the outcome variable. A time-invariant population size offset is still used. For the weekly case fatality rate model, due to delays between COVID-19 diagnosis and official recording of death, we introduced a one-week lag for weekly COVID-19 deaths specific to the USAFacts data. For example, weekly deaths on 12/21/20 for example were shifted back to have an offset for weekly cases on 12/14/20. For case fatality rate models, counties with total case counts of zero were set to one so the log offset would be defined. Additionally, if the number of weekly deaths was greater than the number of matched weekly cases, we set the weekly cases offset to be equal to the corresponding weekly deaths outcome.

Univariable models included a single covariate and its’ interactions with the season categorical variable for *X*_*ij*_, and separate models were fit for each outcome and each covariate. Multivariable models included all covariates and their interactions with season for *X*_*ij*_ and a single model was fit for each outcome. Estimates and confidence intervals for *δ* were used to explore season varying associations between county-level covariates and COVID-19 rates.

All analyses were conducted in R. The following packages were used in formatting data: data.table, dplyr. The following packages were used in formatting results and creating plots: ggplot2, usmap, gridExtra, tidyverse, plyr. The following packages were used in modeling: glmnet, geepack, geeM, lme4, splines, MASS. Code for these analyses is available as described in the code availability section.

## Data Availability

All materials and code for analysis are available on https://github.com/lin-lab/COVID-Health-Disparities.

## Data and code availability

All materials and code for analysis are available on https://github.com/lin-lab.

## Acknowledgments

Resource support was provided by the Cannon cluster supported by the FAS Division of Science, Research Computing Group at Harvard University. This work was funded by NIH grant T32-CA933739 (DL), HD092580 (JTC and BAC), and R35-CA203654-04 (XL).

## Author Contributions

DL, SMG, CQ, and XL contributed to the conception and design of the work and to the acquisition and analysis of the data. All authors contributed to interpreting the data. DL drafted the work. All authors substantively revised the work, approved the submitted work, and agreed to be personally accountable for their own contributions.

## Additional Information

Correspondence and requests for materials should be addressed to Xihong Lin.

## Supplementary Information

### Case Fatality Rate and Infection Fatality Rate Regression

We conducted case fatality rate (CFR) and infection fatality rate (IFR) regression analyses to investigate death rates among infected subjects. CFRs were calculated by dividing the number of deaths by the number of reported cases in each county. CFR regression was performed in a similar way to the cumulative death rate regression by fitting Poisson mixed models except for using an offset for log(total reported cases) instead of log(population size). IFR were calculated by dividing the number of deaths by the number of infected cases in each county.

Since county-specific number of total infected cases were not observed and would likely be underestimated by using the numbers of reported cases, we estimated the county-specific total number of infected cases by dividing the number of total reported cases by a constant ascertainment rate of cases. Since the county-specific testing data and ascertainment rates were not available, we modeled them using state-specific fixed effects and county-specific random effects and allowed the ascertainment rates to vary between states and counties.

To define the CFR model, assume *R*_*ij*_ is the number of reported cases and *μ*_*ij*_ is the mean number of cumulative deaths in county *j* of state *i*. Then the CFR Poisson mixed model can be written as

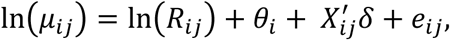

where *θ*_*i*_ is the state effect, *X*_*ij*_ is a vector of covariates, and the *e*_*ij*_ are county-specific random effects to account for overdispersion.

For the IFR model, let *I*_*ij*_ be the unobserved total number of infected cases in county *j* of state *i*. Suppose *a*_*ij*_ is the unobserved ascertainment rate for state *i* and county *j*. The estimated ascertainment rate has been estimated to be between 4% to 20%(30). We have 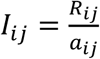. Let *a*_*ij*_ = *c*_*i*_*d*_*ij*_, where *c*_*i*_ is the overall ascertainment rate for state *i* and *d*_*ij*_ is the multiplicative departure of the ascertainment rate of county *j* from the state level acertainment rate *c*_*i*_. Then the IFR Poisson mixed model can be written as

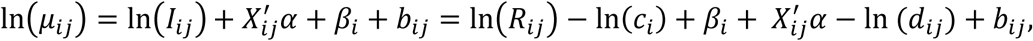

where *β*_*i*_ is the state effect. Write *γ*_*i*_ = −ln(*c*_*i*_) + *β*_*i*_ and *u*_*ij*_ = − ln(*d*_*ij*_) + *b*_*ij*_. Then we

have

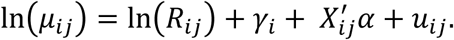

Assuming *u*_*ij*_ to be county-specific random effects following a normal distribution *N*(0, *τ*), it follows that the IFR regression model is identical to the CFR regression model except that the estimated state effects and the county-specific random effects can also be interpreted as capturing the state and county-level ascertainment rates. Therefore, the identical results for CFR and IFR regressions are presented in **Figure 5**. Future work can investigate allowing the county-specific ascertainment rate to be better estimated when county-specific testing data are available.

The CFR/IFR results differed from the cumulative death rate results possibly because of differential underestimation of asymptomatic and mildly asymptomatic cases by race/ethnicity(29), selection bias associated with both the subjects who were tested, e.g., symptomatic subjects were more likely to be tested and to test positive, large fluctuations in the numbers of tests from county to county, and insufficient testing capacity. Additional data collection, such as county-level testing data and race/ethnicity specific case and death counts, is needed to better estimate the number of infected cases. This will allow for more accurate of county-specific ascertainment rates and make IFR analysis results more reliable.

**Fig. S1.**
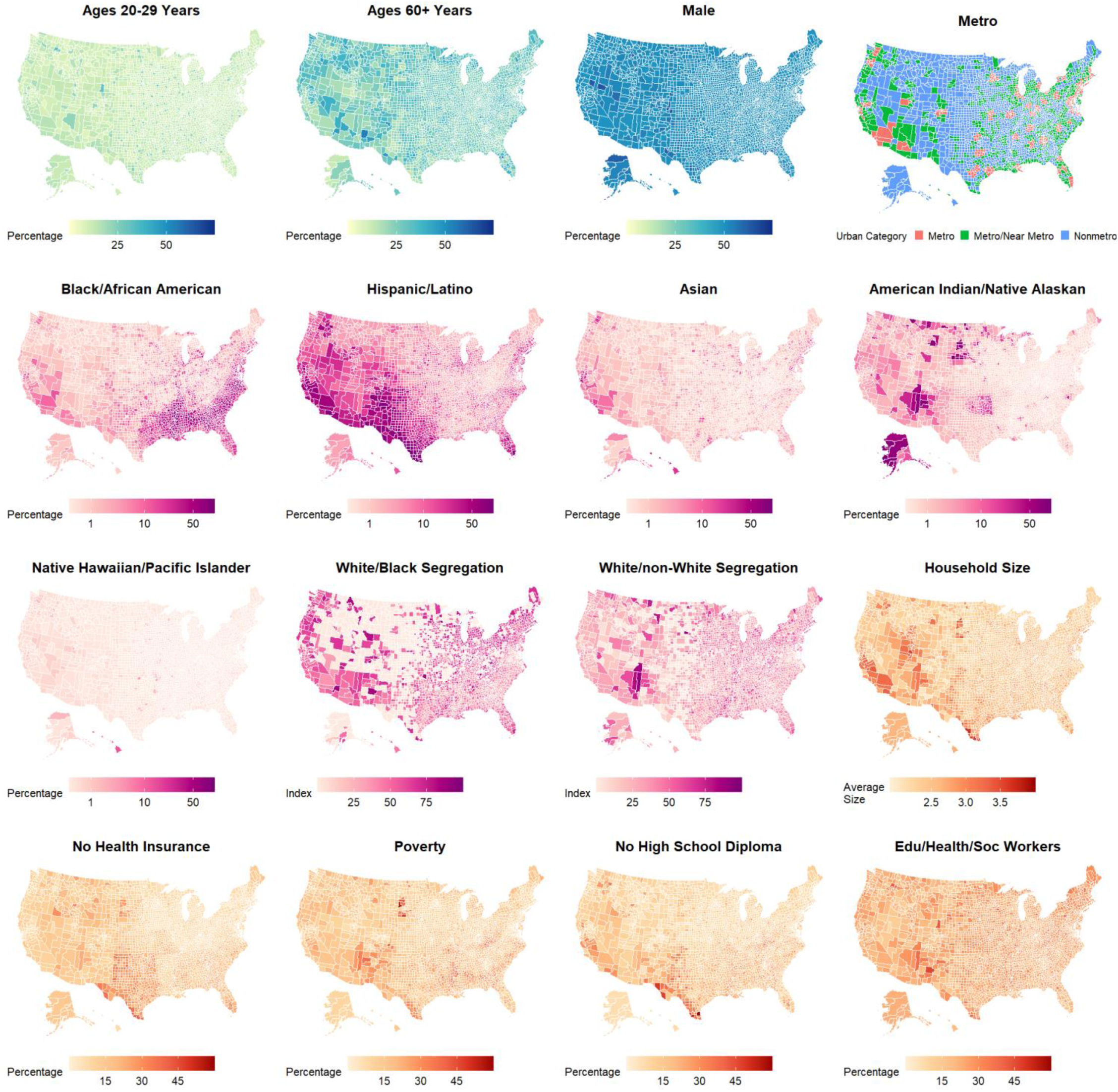
Demographic, racial, and socioeconomic covariate heatmaps. Demographic (yellow, aqua, blue), racial (pink, magenta, purple), and socioeconomic (red, orange) covariate heatmaps.

**Fig. S2.**
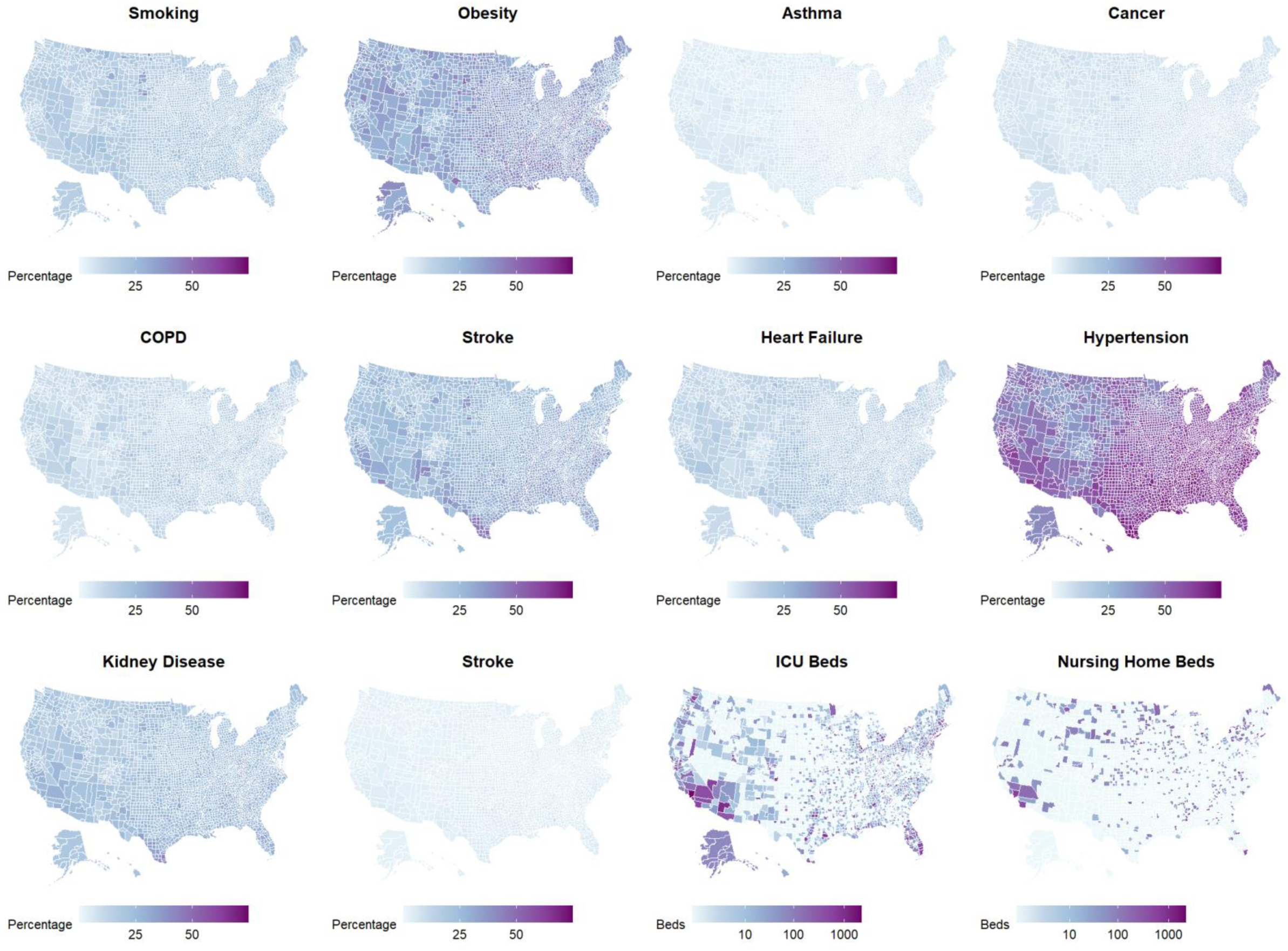
Health covariate heatmaps. Health (white, blue, purple) covariate heatmaps.

**Fig. S3.**
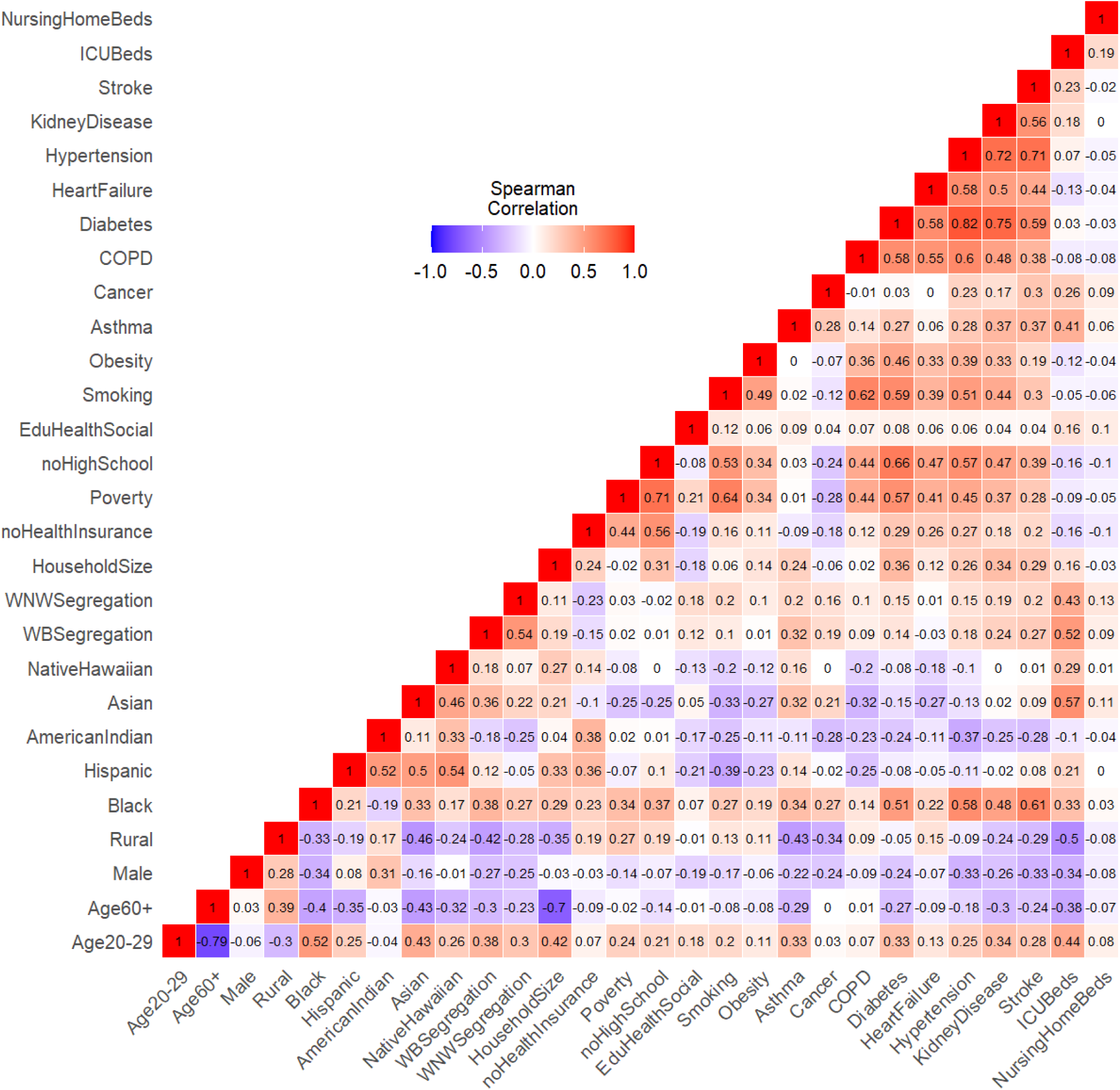
Covariate correlations. Heatmap of Spearman correlations between demographic, racial, socioeconomic, and health covariates.

**Fig. S4.**
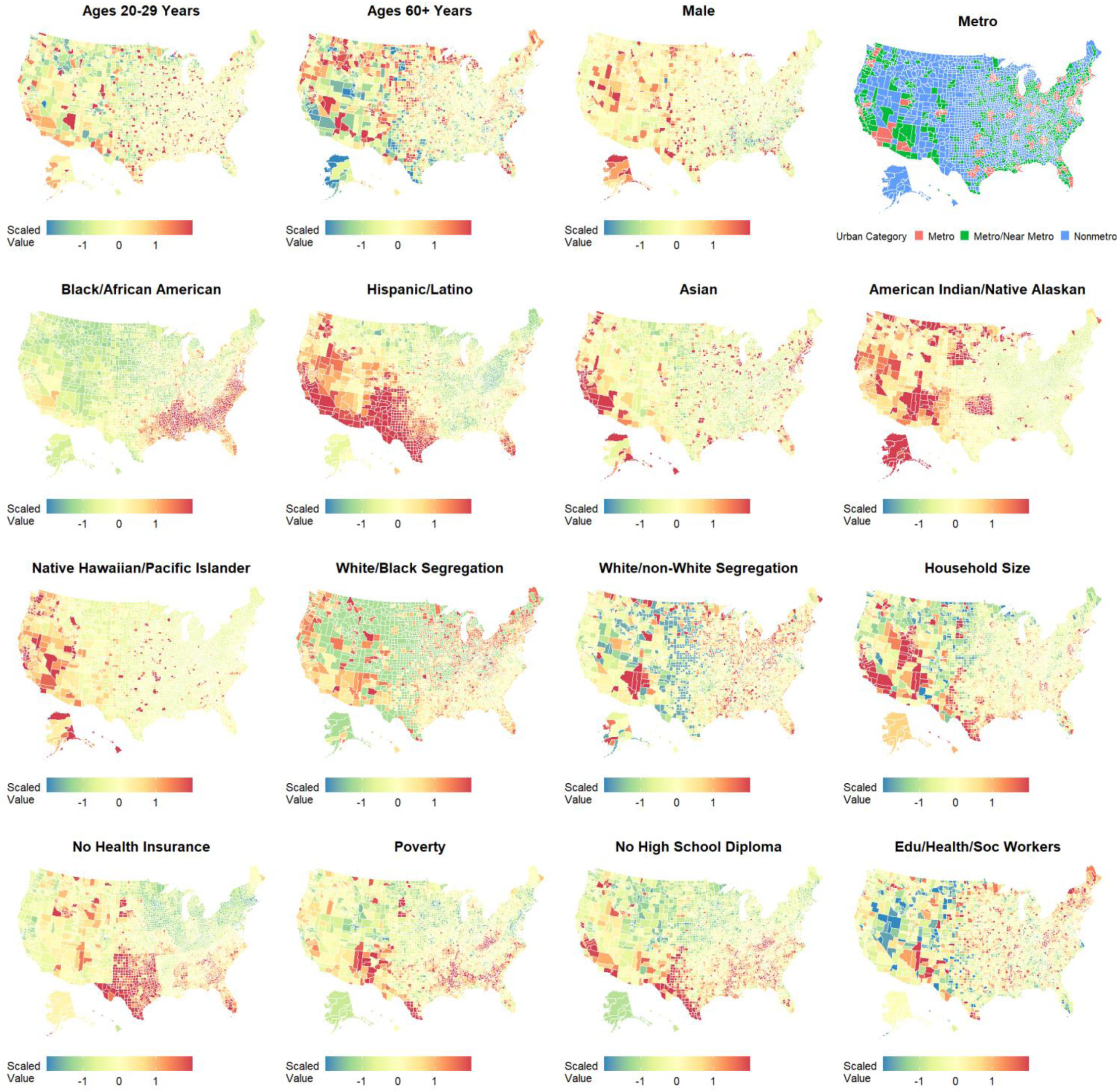
Standardized demographic, racial, and socioeconomic covariate heatmaps. Heatmaps of standardized demographic, racial, and socioeconomic covariates used in regression modeling. For presentation in these heatmaps only, standardized values are cut off at two standard deviations above and below the mean.

**Fig. S5.**
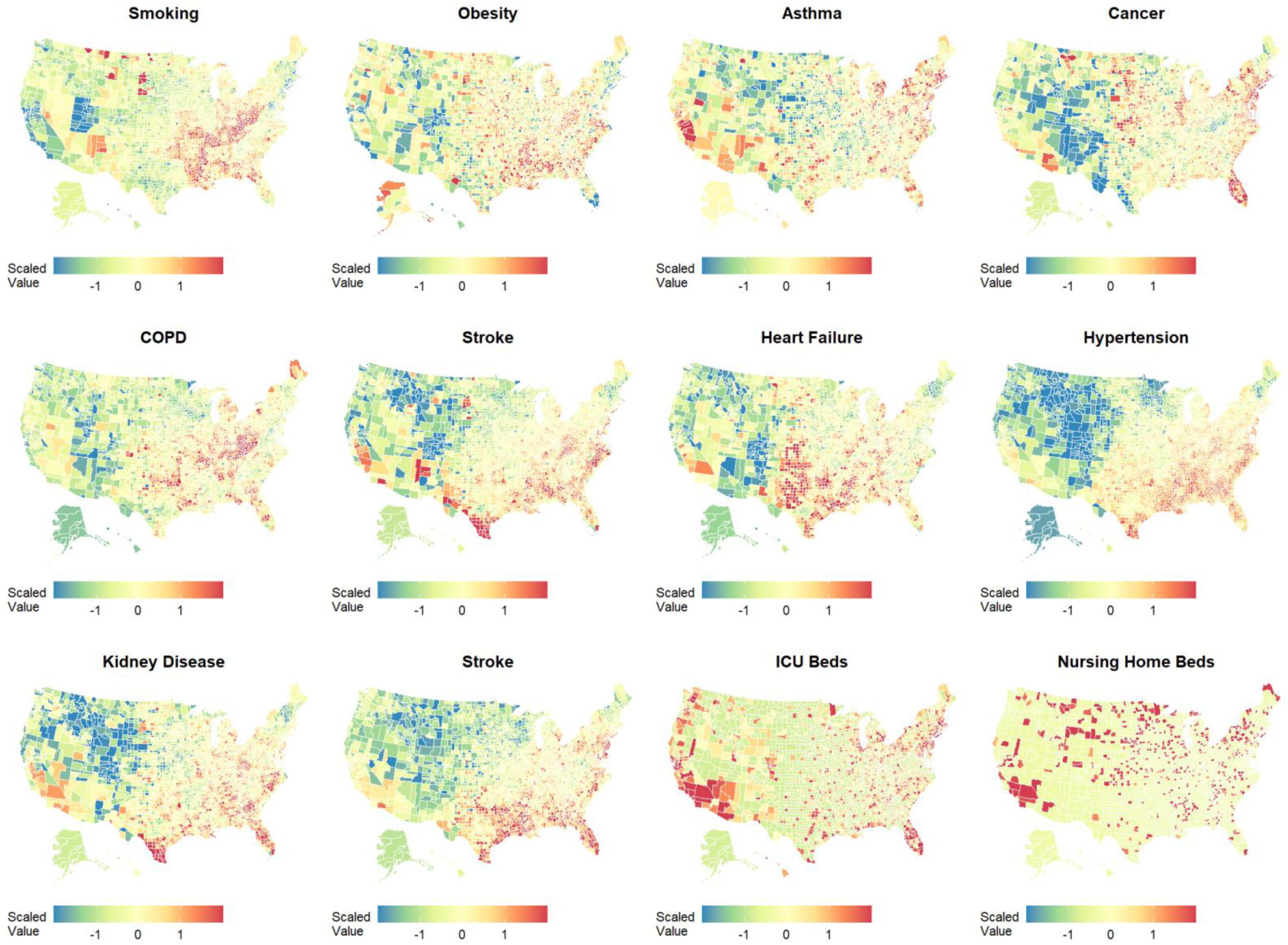
Standardized health covariate heatmaps. Heatmaps of standardized health covariates used in regression modeling. For presentation in these heatmaps only, standardized values are cut off at two standard deviations above and below the mean.

**Fig. S6.**
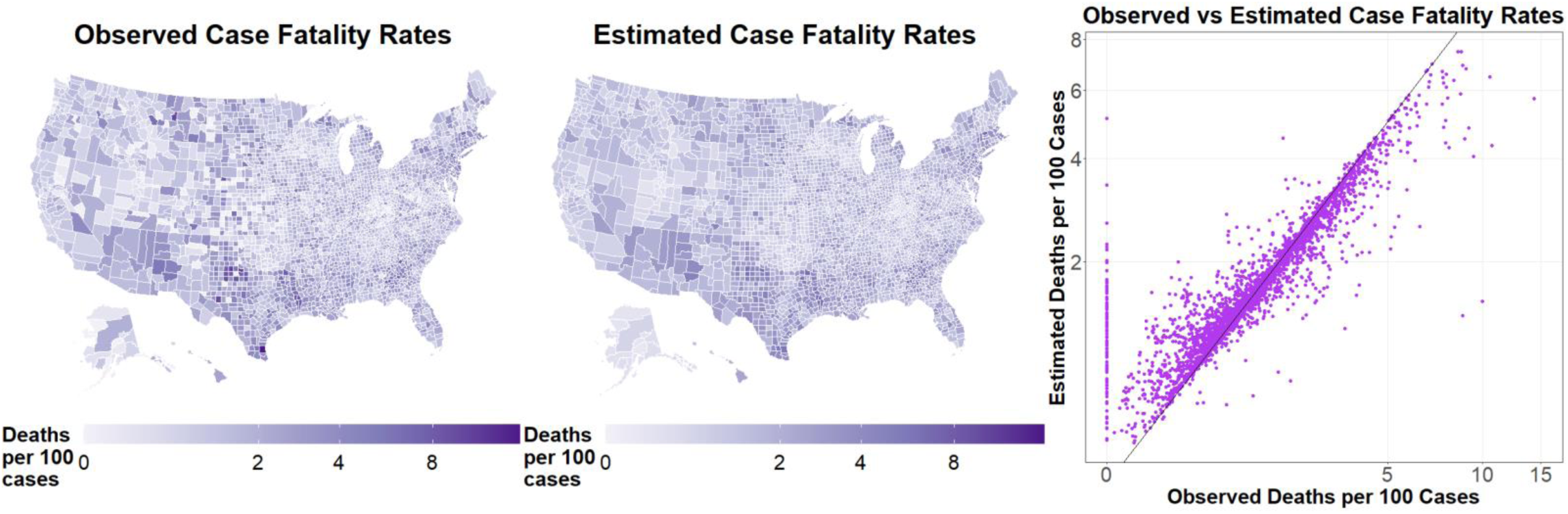
Observed and estimated case, death, and case fatality rates. Observed and multivariable model estimated cumulative case fatality rates through 12/21/2020 for all 3,142 US counties. Left panels are heatmaps of observed rates. Middle panels are heatmaps of estimated rates. Right panels are scatterplots of observed vs estimated rates. The solid line (intercept zero slope one) indicates perfect agreement between observed and estimated rates. Rates are generally well estimated, though 3 counties reported 0 total cases (offsets were set to 1 for modeling) and 111 counties reported 0 total deaths.

**Fig. S7.**
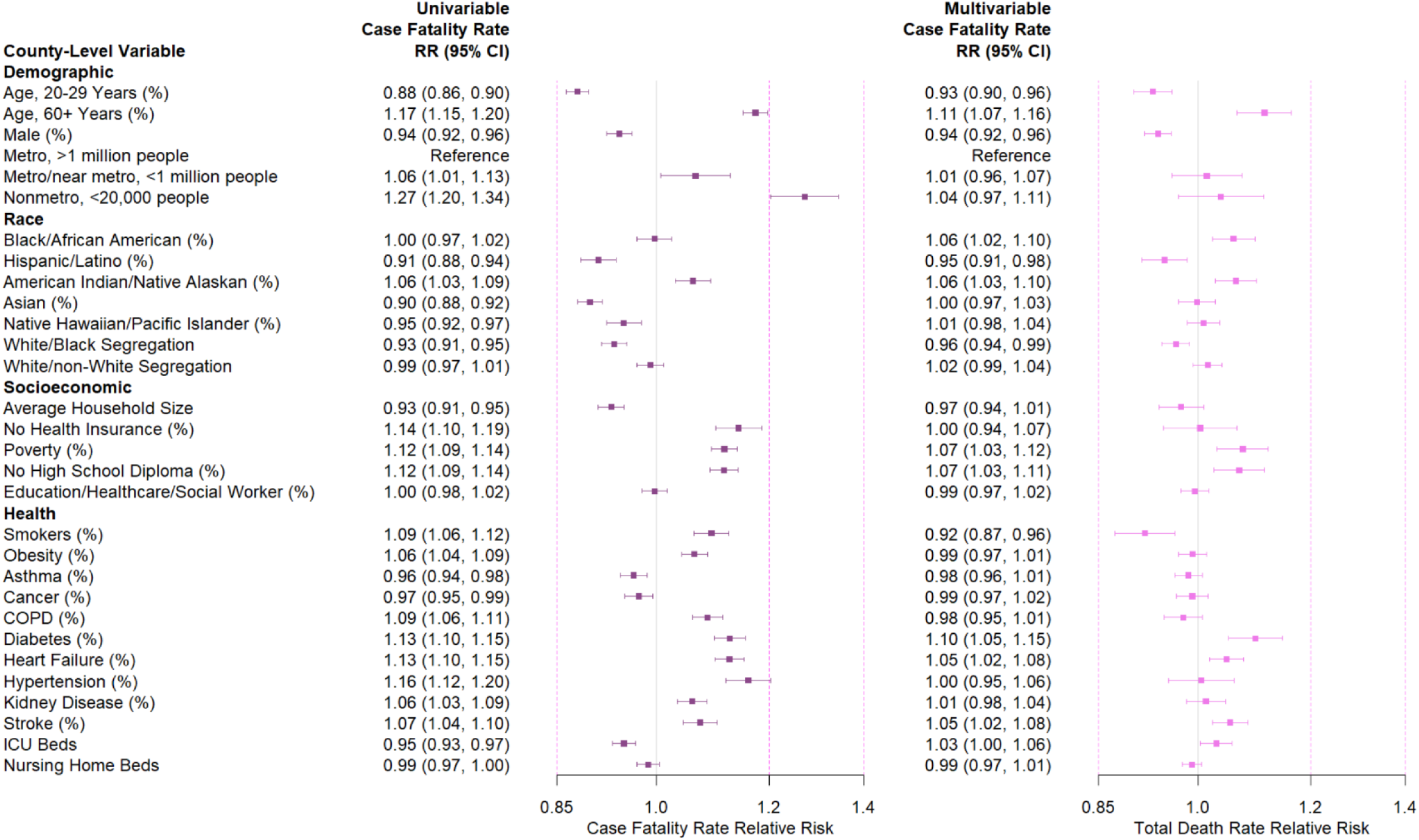
Univariable and multivariable case fatality rate relative risks. Univariable and multivariable relative risks of demographic, socioeconomic, and health comorbidity factors on cumulative COVID-19 case fatality rates through 12/21/20 additionally adjust for state fixed effects and county random effects. Boxes are point estimates and error bars mark 95% confidence intervals. Relative risks are for a one standard deviation increase in a variable (see **Table S1**), except for the metro/nonmetro categorical variable.

**Fig. S8.**
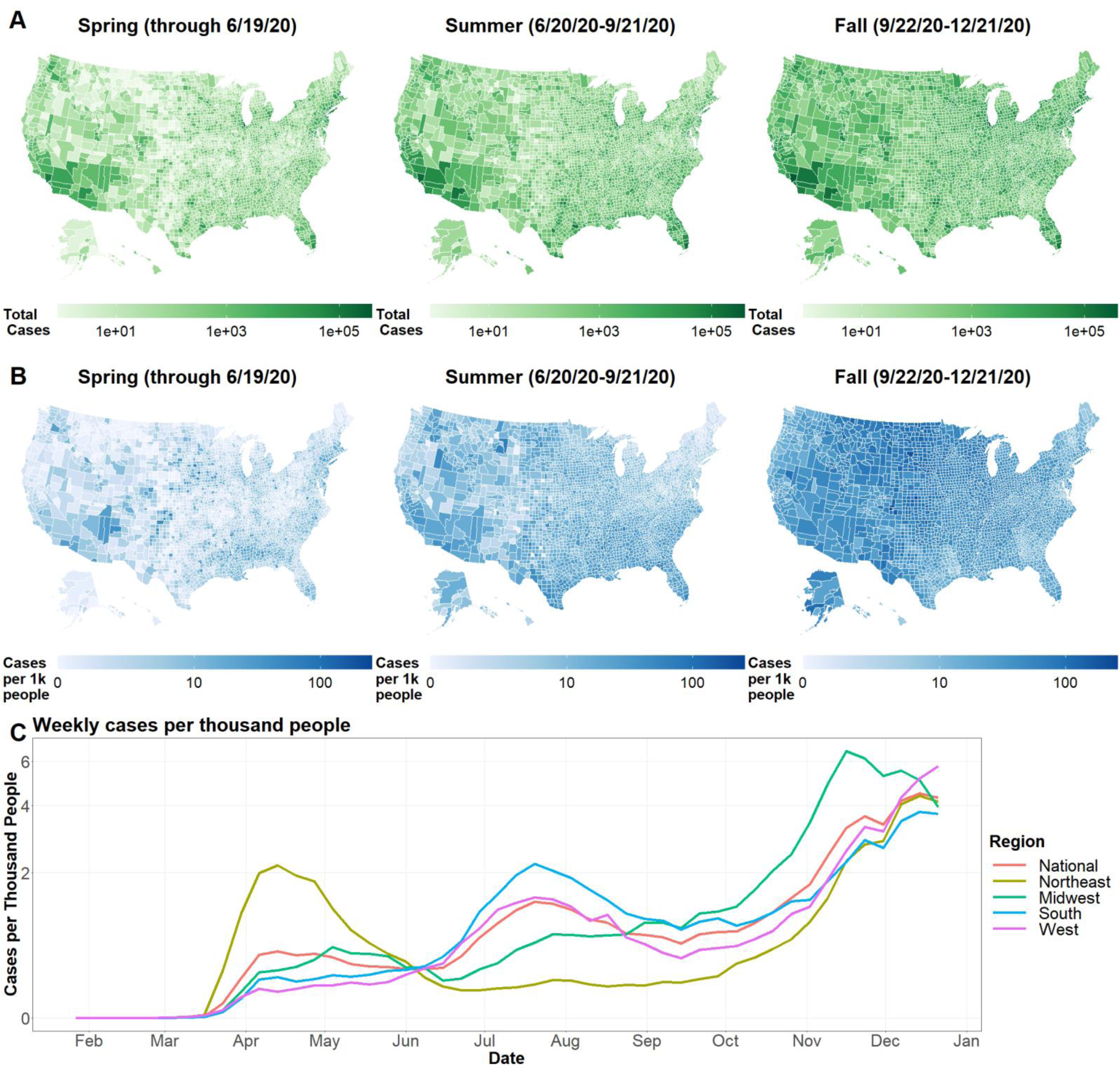
Weekly case rates. (A) Heatmaps of county total cases by season. (B) Heatmaps of county total case rates by season. (C) Line plots of weekly case rates nationally and by US Census region.

**Fig. S9.**
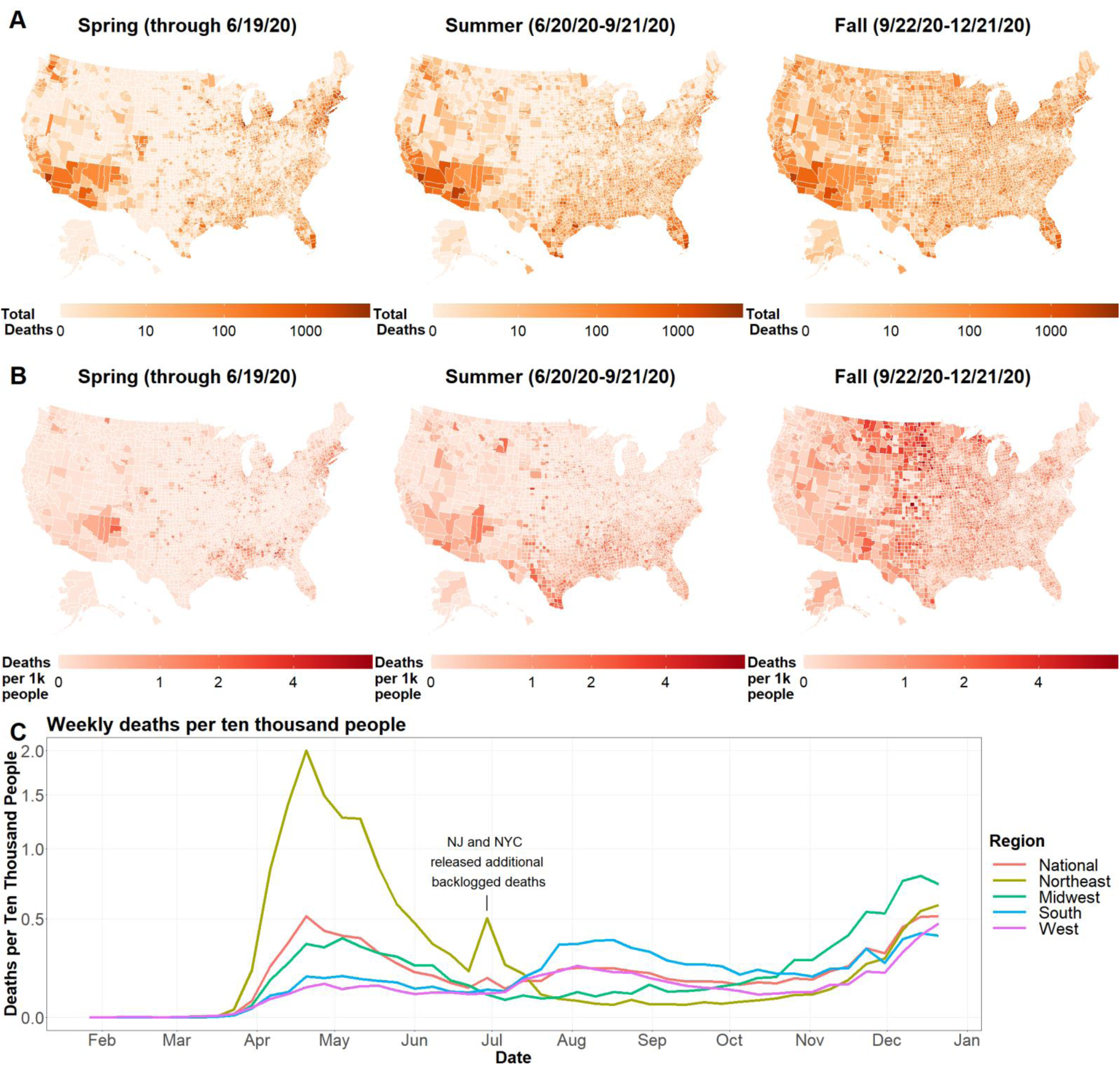
Weekly death rates. (A) Heatmaps of county total deaths by season. (B) Heatmaps of county total death rates by season. (C) Line plots of weekly death rates nationally and by US Census region.

**Fig. S10.**
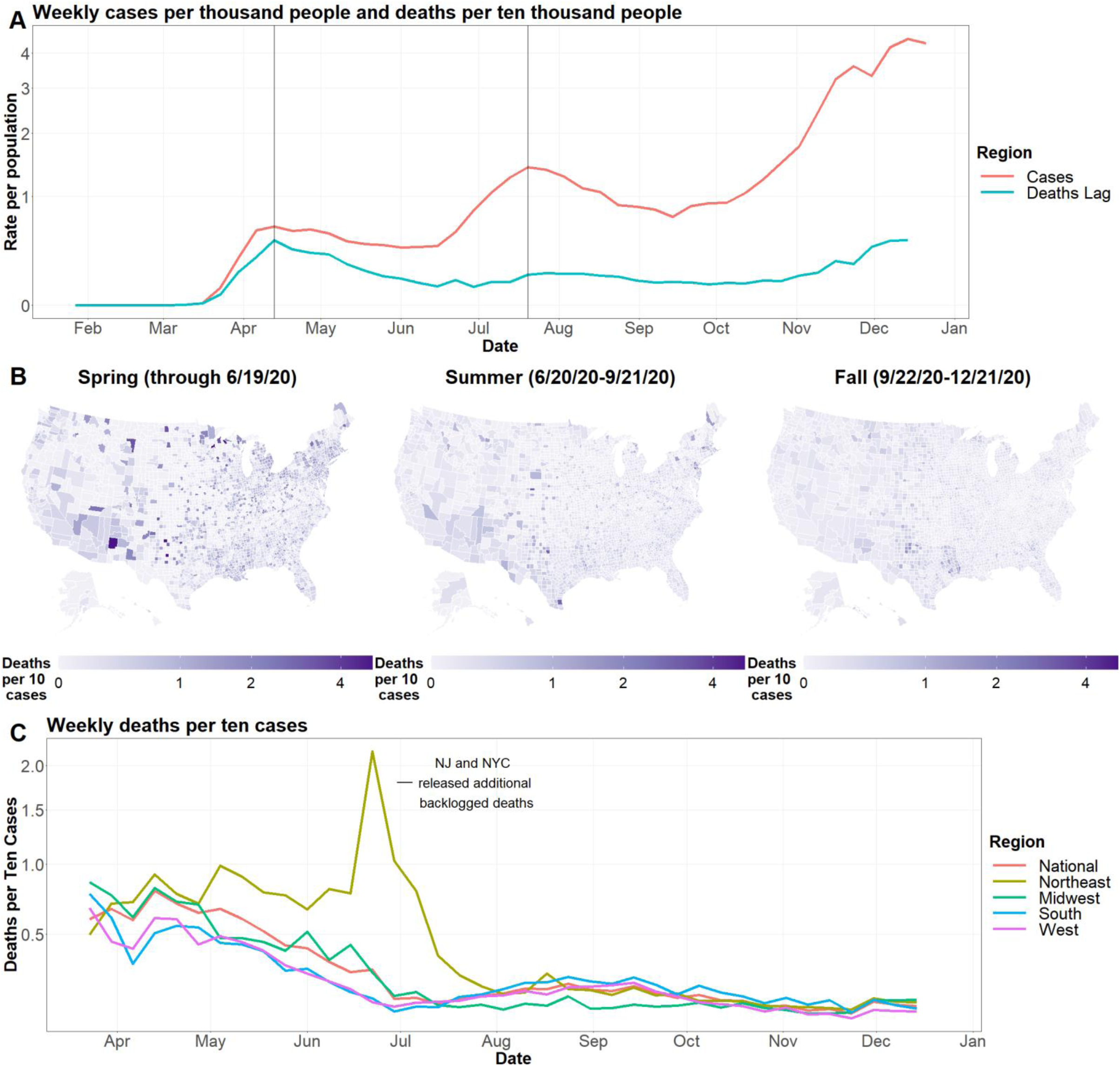
Weekly case fatality rates. (A) Line plots of US national weekly case rates and death rates lagged by one week. Solid lines mark similar peaks between weekly case rates and lagged death rates. (B) Heatmaps of county case fatality rates by season. (C) Line plots of weekly case fatality rates nationally and by US Census region.

**Table S1.** List of county-level variables, transformations, and sources.

| Variable Name | Date | Transformation | Source and Description |
| --- | --- | --- | --- |
| **Outcomes** |  |  |  |
| Cumulative COVID-19 Cases | 12/21/20 |  | USA Facts |
| Cumulative COVID-19 Deaths | 12/21/20 |  | USA Facts |
| **Demographics** |  |  |  |
| Population Size | 2019 | log offset | US Census Bureau |
| Age, 20-29 years (%) | 2019 |  | US Census Bureau |
| Age, 60+ years (%) | 2018 |  | US Census Bureau |
| Male (%) | 2018 |  | US Census Bureau |
| Rural Urban Continuum Code | 2013 |  | US Department of Agriculture |
| **Racial** |  |  |  |
| Black/African American (%) | 2019 | log(x+1) | US Census Bureau |
| Asian (%) | 2019 | log(x+1) | US Census Bureau |
| Hispanic/Latino (%) | 2019 | log(x+1) | US Census Bureau |
| American Indian/Native Alaskan (%) | 2019 | log(x+1) | US Census Bureau |
| Native Hawaiian/Pacific Islander (%) | 2019 | log(x+1) | US Census Bureau |
| White Black Segregation Index | 2014-2018 |  | County Health Rankings & Roadmaps |
| White non-White Segregation Index | 2014-2018 |  | County Health Rankings & Roadmaps |
| **Socioeconomic** |  |  |  |
| Average Household Size | 2010 |  | Area Health Resources Files |
| No Health Insurance, 18-64 years (%) | 2017 |  | Area Health Resources Files |
| Poverty (%) | 2017 |  | Area Health Resources Files |
| No High School Diploma, 25+ years (%) | 2013-17 |  | Area Health Resources Files |
| Education, Health Care, Social Assistance Workers (%) | 2013-17 |  | Area Health Resources Files |
| **Health** |  |  |  |
| Smokers (%) | 2017 |  | County Health Rankings & Roadmaps |
| Obesity (%) | 2017 |  | County Health Rankings & Roadmaps |
| Asthma (%) | 2017 |  | County Health Rankings & Roadmaps |
| Cancer (%) | 2017 |  | County Health Rankings & Roadmaps |
| COPD (%) | 2017 |  | County Health Rankings & Roadmaps |
| Diabetes (%) | 2017 |  | County Health Rankings & Roadmaps |
| Heart Failure (%) | 2017 |  | County Health Rankings & Roadmaps |
| Hypertension (%) | 2017 |  | County Health Rankings & Roadmaps |
| Kidney Disease (%) | 2017 |  | County Health Rankings & Roadmaps |
| Stroke (%) | 2017 |  | County Health Rankings & Roadmaps |
| ICU Beds | 2017 | $\log(x+1)$ | Kaiser Health News |
| Nursing Home Beds | 2017 | $\log(x+1)$ | Kaiser Health News |

**Table S2.**
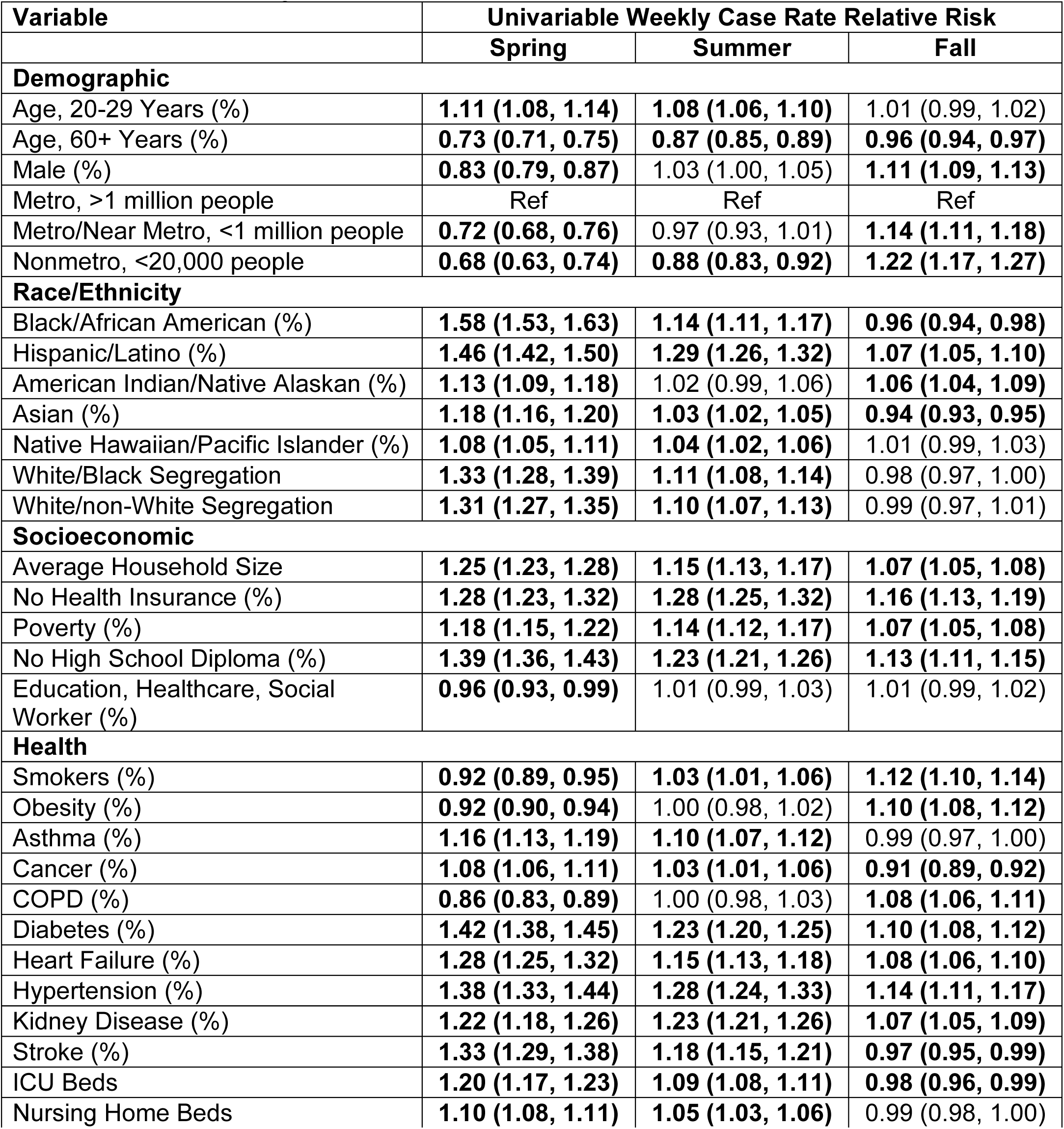
Univariable weekly case rates. Relative risks of county-level variables on weekly case rates (40 repeated measurements per county) by season from 3/23/20-12/21/20. Each row is a separate model and controls for state effects, US census region-specific time varying trends, and additional county overdispersion. Parentheses indicate 95% confidence intervals. Bold indicates confidence interval does not contain 1. Relative risks are for a one standard deviation increase in a variable, except for the metro/nonmetro categorical variable.

| Variable | Univariable Weekly Case Rate Relative Risk |  |  |
| --- | --- | --- | --- |
|  | Spring | Summer | Fall |
| **Demographic** |  |  |  |
| Age, 20-29 Years (%) | **1.11 (1.08, 1.14)** | **1.08 (1.06, 1.10)** | 1.01 (0.99, 1.02) |
| Age, 60+ Years (%) | **0.73 (0.71, 0.75)** | **0.87 (0.85, 0.89)** | **0.96 (0.94, 0.97)** |
| Male (%) | **0.83 (0.79, 0.87)** | 1.03 (1.00, 1.05) | **1.11 (1.09, 1.13)** |
| Metro, >1 million people | Ref | Ref | Ref |
| Metro/Near Metro, <1 million people | **0.72 (0.68, 0.76)** | 0.97 (0.93, 1.01) | **1.14 (1.11, 1.18)** |
| Nonmetro, <20,000 people | **0.68 (0.63, 0.74)** | **0.88 (0.83, 0.92)** | **1.22 (1.17, 1.27)** |
| **Race/Ethnicity** |  |  |  |
| Black/African American (%) | **1.58 (1.53, 1.63)** | **1.14 (1.11, 1.17)** | **0.96 (0.94, 0.98)** |
| Hispanic/Latino (%) | **1.46 (1.42, 1.50)** | **1.29 (1.26, 1.32)** | **1.07 (1.05, 1.10)** |
| American Indian/Native Alaskan (%) | **1.13 (1.09, 1.18)** | 1.02 (0.99, 1.06) | **1.06 (1.04, 1.09)** |
| Asian (%) | **1.18 (1.16, 1.20)** | **1.03 (1.02, 1.05)** | **0.94 (0.93, 0.95)** |
| Native Hawaiian/Pacific Islander (%) | **1.08 (1.05, 1.11)** | **1.04 (1.02, 1.06)** | 1.01 (0.99, 1.03) |
| White/Black Segregation | **1.33 (1.28, 1.39)** | **1.11 (1.08, 1.14)** | 0.98 (0.97, 1.00) |
| White/non-White Segregation | **1.31 (1.27, 1.35)** | **1.10 (1.07, 1.13)** | 0.99 (0.97, 1.01) |
| **Socioeconomic** |  |  |  |
| Average Household Size | **1.25 (1.23, 1.28)** | **1.15 (1.13, 1.17)** | **1.07 (1.05, 1.08)** |
| No Health Insurance (%) | **1.28 (1.23, 1.32)** | **1.28 (1.25, 1.32)** | **1.16 (1.13, 1.19)** |
| Poverty (%) | **1.18 (1.15, 1.22)** | **1.14 (1.12, 1.17)** | **1.07 (1.05, 1.08)** |
| No High School Diploma (%) | **1.39 (1.36, 1.43)** | **1.23 (1.21, 1.26)** | **1.13 (1.11, 1.15)** |
| Education, Healthcare, Social Worker (%) | **0.96 (0.93, 0.99)** | 1.01 (0.99, 1.03) | 1.01 (0.99, 1.02) |
| **Health** |  |  |  |
| Smokers (%) | **0.92 (0.89, 0.95)** | **1.03 (1.01, 1.06)** | **1.12 (1.10, 1.14)** |
| Obesity (%) | **0.92 (0.90, 0.94)** | 1.00 (0.98, 1.02) | **1.10 (1.08, 1.12)** |
| Asthma (%) | **1.16 (1.13, 1.19)** | **1.10 (1.07, 1.12)** | 0.99 (0.97, 1.00) |
| Cancer (%) | **1.08 (1.06, 1.11)** | **1.03 (1.01, 1.06)** | **0.91 (0.89, 0.92)** |
| COPD (%) | **0.86 (0.83, 0.89)** | 1.00 (0.98, 1.03) | **1.08 (1.06, 1.11)** |
| Diabetes (%) | **1.42 (1.38, 1.45)** | **1.23 (1.20, 1.25)** | **1.10 (1.08, 1.12)** |
| Heart Failure (%) | **1.28 (1.25, 1.32)** | **1.15 (1.13, 1.18)** | **1.08 (1.06, 1.10)** |
| Hypertension (%) | **1.38 (1.33, 1.44)** | **1.28 (1.24, 1.33)** | **1.14 (1.11, 1.17)** |
| Kidney Disease (%) | **1.22 (1.18, 1.26)** | **1.23 (1.21, 1.26)** | **1.07 (1.05, 1.09)** |
| Stroke (%) | **1.33 (1.29, 1.38)** | **1.18 (1.15, 1.21)** | **0.97 (0.95, 0.99)** |
| ICU Beds | **1.20 (1.17, 1.23)** | **1.09 (1.08, 1.11)** | **0.98 (0.96, 0.99)** |
| Nursing Home Beds | **1.10 (1.08, 1.11)** | **1.05 (1.03, 1.06)** | 0.99 (0.98, 1.00) |

**Table S3.** Multivariable weekly case rates. Relative risks of county-level variables on weekly case rates (40 repeated measurements per county) by season from 3/23/20-12/21/20. All results are from a single model that controls for state effects, US census region-specific time varying trends, and additional county overdispersion. Parentheses indicate 95% confidence intervals. Bold indicates confidence interval does not contain 1. Relative risks are for a one standard deviation increase in a variable, except for the metro/nonmetro categorical variable.

| Variable | Multivariable Weekly Case Rate Relative Risk |  |  |
| --- | --- | --- | --- |
|  | Spring | Summer | Fall |
| **Demographic** |  |  |  |
| Age, 20-29 Years (%) | 1.00 (0.95, 1.05) | 1.01 (0.98, 1.04) | 0.99 (0.97, 1.02) |
| Age, 60+ Years (%) | **0.92 (0.86, 0.97)** | **0.94 (0.90, 0.98)** | **0.93 (0.90, 0.96)** |
| Male (%) | **1.05 (1.01, 1.10)** | **1.09 (1.06, 1.12)** | **1.04 (1.02, 1.07)** |
| Metro, >1 million people | Ref | Ref | Ref |
| Metro/Near Metro, <1 million people | 0.94 (0.88, 1.01) | 1.05 (1.00, 1.10) | **1.06 (1.02, 1.10)** |
| Nonmetro, <20,000 people | 1.01 (0.89, 1.14) | 1.04 (0.97, 1.13) | **1.09 (1.03, 1.14)** |
| **Race/Ethnicity** |  |  |  |
| Black/African American (%) | **1.48 (1.41, 1.56)** | 0.97 (0.94, 1.00) | **0.96 (0.93, 0.98)** |
| Hispanic/Latino (%) | **1.18 (1.13, 1.24)** | **1.17 (1.14, 1.21)** | **1.04 (1.01, 1.07)** |
| American Indian/Native Alaskan (%) | **1.13 (1.08, 1.19)** | **0.95 (0.92, 0.98)** | 1.01 (0.99, 1.04) |
| Asian (%) | 0.97 (0.94, 1.00) | **0.97 (0.95, 0.99)** | **0.93 (0.91, 0.95)** |
| Native Hawaiian/Pacific Islander (%) | 1.00 (0.97, 1.03) | 1.02 (1.00, 1.04) | 0.99 (0.97, 1.01) |
| White/Black Segregation | **1.12 (1.06, 1.18)** | 1.01 (0.98, 1.04) | 1.00 (0.98, 1.02) |
| White/non-White Segregation | **1.15 (1.10, 1.20)** | **1.07 (1.04, 1.11)** | 1.02 (1.00, 1.04) |
| **Socioeconomic** |  |  |  |
| Average Household Size | 1.04 (1.00, 1.08) | 1.03 (1.00, 1.06) | 1.01 (0.99, 1.04) |
| No Health Insurance (%) | **0.86 (0.82, 0.91)** | 0.99 (0.95, 1.04) | **0.95 (0.91, 0.99)** |
| Poverty (%) | 0.95 (0.89, 1.01) | **1.06 (1.02, 1.11)** | **0.95 (0.92, 0.98)** |
| No High School Diploma (%) | **1.37 (1.29, 1.45)** | **1.10 (1.06, 1.15)** | **1.12 (1.08, 1.15)** |
| Education, Healthcare, Social Worker (%) | 0.98 (0.95, 1.02) | **1.06 (1.03, 1.08)** | 1.02 (1.00, 1.04) |
| **Health** |  |  |  |
| Smokers (%) | **0.91 (0.86, 0.96)** | 1.01 (0.97, 1.05) | 0.99 (0.96, 1.03) |
| Obesity (%) | **0.95 (0.92, 0.99)** | 1.01 (0.98, 1.03) | 1.00 (0.98, 1.02) |
| Asthma (%) | **0.95 (0.92, 0.99)** | 1.00 (0.98, 1.02) | 0.99 (0.97, 1.00) |
| Cancer (%) | **1.08 (1.03, 1.12)** | **1.07 (1.04, 1.10)** | **0.96 (0.94, 0.98)** |
| COPD (%) | **0.90 (0.86, 0.95)** | **0.89 (0.86, 0.92)** | 1.00 (0.97, 1.02) |
| Diabetes (%) | **1.15 (1.10, 1.21)** | **0.94 (0.90, 0.98)** | 0.99 (0.96, 1.02) |
| Heart Failure (%) | 0.97 (0.93, 1.02) | 1.02 (0.99, 1.05) | **1.06 (1.04, 1.09)** |
| Hypertension (%) | **1.16 (1.07, 1.25)** | **1.17 (1.11, 1.24)** | **1.10 (1.05, 1.15)** |
| Kidney Disease (%) | **0.83 (0.79, 0.88)** | **1.05 (1.01, 1.08)** | 1.00 (0.97, 1.02) |
| Stroke (%) | 0.99 (0.94, 1.03) | 1.03 (1.00, 1.07) | **0.95 (0.93, 0.97)** |
| ICU Beds | **0.92 (0.89, 0.96)** | 0.99 (0.96, 1.01) | **1.09 (1.07, 1.11)** |
| Nursing Home Beds | **0.97 (0.95, 0.98)** | **1.02 (1.01, 1.03)** | 1.00 (0.99, 1.01) |

**Table S4.** Univariable weekly death rates. Relative risks of county-level variables on weekly death rates (40 repeated measurements per county) by season from 3/23/20-12/21/20. Each row is a separate model and controls for state effects, US census region-specific time varying trends, and additional county overdispersion. Parentheses indicate 95% confidence intervals. Bold indicates confidence interval does not contain 1. Relative risks are for a one standard deviation increase in a variable, except for the metro/nonmetro categorical variable.

| Variable | Univariable Weekly Death Rate Relative Risk |  |  |
| --- | --- | --- | --- |
|  | Spring | Summer | Fall |
| **Demographic** |  |  |  |
| Age, 20-29 Years (%) | **1.08 (1.05, 1.11)** | **0.94 (0.92, 0.97)** | **0.83 (0.81, 0.86)** |
| Age, 60+ Years (%) | **0.77 (0.74, 0.80)** | 1.01 (0.97, 1.04) | **1.27 (1.24, 1.30)** |
| Male (%) | **0.45 (0.41, 0.48)** | **0.93 (0.88, 0.98)** | **1.21 (1.17, 1.26)** |
| Metro, >1 million people | Ref | Ref | Ref |
| Metro/Near Metro, <1 million people | **0.59 (0.55, 0.63)** | **1.08 (1.01, 1.15)** | **1.80 (1.69, 1.91)** |
| Nonmetro, <20,000 people | **0.53 (0.48, 0.58)** | **1.16 (1.07, 1.25)** | **2.64 (2.47, 2.82)** |
| **Race/Ethnicity** |  |  |  |
| Black/African American (%) | **2.00 (1.92, 2.08)** | **1.10 (1.06, 1.14)** | **0.76 (0.74, 0.79)** |
| Hispanic/Latino (%) | **1.31 (1.26, 1.36)** | **1.22 (1.18, 1.27)** | **0.85 (0.82, 0.88)** |
| American Indian/Native Alaskan (%) | **1.17 (1.12, 1.23)** | **1.11 (1.06, 1.17)** | **1.15 (1.10, 1.20)** |
| Asian (%) | **1.18 (1.16, 1.21)** | **0.94 (0.92, 0.96)** | **0.74 (0.72, 0.75)** |
| Native Hawaiian/Pacific Islander (%) | 1.03 (0.99, 1.07) | 1.00 (0.97, 1.04) | **0.94 (0.91, 0.98)** |
| White/Black Segregation | **1.48 (1.42, 1.55)** | **1.06 (1.02, 1.10)** | **0.79 (0.76, 0.81)** |
| White/non-White Segregation | **1.53 (1.47, 1.59)** | **1.09 (1.05, 1.13)** | **0.86 (0.83, 0.89)** |
| **Socioeconomic** |  |  |  |
| Average Household Size | **1.15 (1.12, 1.19)** | **1.15 (1.12, 1.19)** | **0.90 (0.87, 0.92)** |
| No Health Insurance (%) | **1.32 (1.26, 1.39)** | **1.60 (1.53, 1.67)** | **1.33 (1.28, 1.39)** |
| Poverty (%) | **1.32 (1.28, 1.36)** | **1.32 (1.28, 1.36)** | **1.17 (1.14, 1.21)** |
| No High School Diploma (%) | **1.45 (1.41, 1.50)** | **1.49 (1.45, 1.53)** | **1.18 (1.15, 1.21)** |
| Education, Healthcare, Social Worker (%) | **0.96 (0.92, 0.99)** | 1.02 (0.99, 1.06) | **1.06 (1.03, 1.09)** |
| **Health** |  |  |  |
| Smokers (%) | **0.99 (0.95, 1.03)** | **1.15 (1.11, 1.19)** | **1.39 (1.34, 1.44)** |
| Obesity (%) | **0.86 (0.83, 0.89)** | **1.11 (1.08, 1.14)** | **1.33 (1.29, 1.36)** |
| Asthma (%) | **1.22 (1.18, 1.26)** | **1.06 (1.02, 1.09)** | **0.83 (0.80, 0.85)** |
| Cancer (%) | **1.12 (1.08, 1.16)** | **0.94 (0.91, 0.98)** | **0.78 (0.76, 0.80)** |
| COPD (%) | **0.88 (0.85, 0.92)** | **1.13 (1.10, 1.18)** | **1.31 (1.27, 1.35)** |
| Diabetes (%) | **1.62 (1.57, 1.67)** | **1.49 (1.44, 1.53)** | **1.11 (1.08, 1.14)** |
| Heart Failure (%) | **1.53 (1.48, 1.58)** | **1.34 (1.30, 1.39)** | **1.12 (1.09, 1.16)** |
| Hypertension (%) | **1.61 (1.52, 1.69)** | **1.70 (1.62, 1.79)** | **1.22 (1.17, 1.28)** |
| Kidney Disease (%) | **1.35 (1.30, 1.40)** | **1.43 (1.38, 1.48)** | 1.02 (0.99, 1.05) |
| Stroke (%) | **1.57 (1.51, 1.64)** | **1.31 (1.26, 1.36)** | **0.85 (0.82, 0.88)** |
| ICU Beds | **1.35 (1.32, 1.39)** | **1.07 (1.04, 1.10)** | **0.81 (0.79, 0.83)** |
| Nursing Home Beds | **1.15 (1.12, 1.17)** | **1.04 (1.02, 1.06)** | **0.90 (0.88, 0.92)** |

**Table S5.** Multivariable weekly death rates. Relative risks of county-level variables on weekly death rates (40 repeated measurements per county) by season from 3/23/20-12/21/20. All results are from a single model that controls for state effects, US census region-specific time varying trends, and additional county overdispersion. Parentheses indicate 95% confidence intervals. Bold indicates confidence interval does not contain 1. Relative risks are for a one standard deviation increase in a variable, except for the metro/nonmetro categorical variable.

| Variable | Multivariable Weekly Death Rate Relative Risk |  |  |
| --- | --- | --- | --- |
|  | Spring | Summer | Fall |
| **Demographic** |  |  |  |
| Age, 20-29 Years (%) | 0.97 (0.92, 1.03) | **0.88 (0.84, 0.92)** | **0.94 (0.91, 0.98)** |
| Age, 60+ Years (%) | **1.13 (1.06, 1.21)** | 1.05 (0.99, 1.12) | 1.00 (0.95, 1.06) |
| Male (%) | **0.80 (0.75, 0.85)** | 1.01 (0.97, 1.05) | 1.04 (1.00, 1.07) |
| Metro, >1 million people | Ref | Ref | Ref |
| Metro/Near Metro, <1 million people | **0.90 (0.84, 0.97)** | 1.01 (0.94, 1.08) | **1.23 (1.16, 1.31)** |
| Nonmetro, <20,000 people | 0.91 (0.80, 1.05) | 0.99 (0.89, 1.11) | **1.35 (1.24, 1.47)** |
| **Race/Ethnicity** |  |  |  |
| Black/African American (%) | **1.89 (1.78, 2.00)** | 1.00 (0.95, 1.05) | **0.88 (0.84, 0.92)** |
| Hispanic/Latino (%) | 0.97 (0.92, 1.03) | **1.10 (1.05, 1.16)** | 1.00 (0.96, 1.04) |
| American Indian/Native Alaskan (%) | **1.34 (1.26, 1.41)** | 1.03 (0.98, 1.08) | 1.02 (0.98, 1.06) |
| Asian (%) | 0.96 (0.93, 1.00) | 1.01 (0.97, 1.05) | **0.89 (0.87, 0.92)** |
| Native Hawaiian/Pacific Islander (%) | **0.91 (0.87, 0.95)** | 1.03 (1.00, 1.06) | 1.03 (1.00, 1.06) |
| White/Black Segregation | **1.07 (1.01, 1.13)** | 0.98 (0.94, 1.03) | **0.96 (0.93, 0.99)** |
| White/non-White Segregation | **1.09 (1.03, 1.15)** | **1.09 (1.04, 1.14)** | 1.03 (1.00, 1.07) |
| **Socioeconomic** |  |  |  |
| Average Household Size | **1.13 (1.08, 1.18)** | 0.99 (0.95, 1.04) | **0.91 (0.88, 0.95)** |
| No Health Insurance (%) | **0.72 (0.67, 0.77)** | 1.04 (0.97, 1.11) | 0.99 (0.93, 1.06) |
| Poverty (%) | **1.16 (1.08, 1.25)** | **1.07 (1.01, 1.14)** | 0.96 (0.91, 1.02) |
| No High School Diploma (%) | **1.20 (1.12, 1.29)** | **1.25 (1.18, 1.33)** | **1.15 (1.09, 1.22)** |
| Education, Healthcare, Social Worker (%) | **0.82 (0.79, 0.86)** | **1.11 (1.07, 1.15)** | **1.06 (1.03, 1.09)** |
| **Health** |  |  |  |
| Smokers (%) | **0.84 (0.79, 0.90)** | **0.97 (0.91, 1.03)** | 0.97 (0.92, 1.03) |
| Obesity (%) | **0.81 (0.78, 0.85)** | 1.01 (0.97, 1.05) | **1.04 (1.01, 1.07)** |
| Asthma (%) | 0.96 (0.93, 1.00) | 0.98 (0.95, 1.02) | 0.98 (0.96, 1.01) |
| Cancer (%) | 1.01 (0.96, 1.05) | 1.01 (0.97, 1.05) | **0.94 (0.91, 0.97)** |
| COPD (%) | **0.93 (0.88, 0.99)** | **0.87 (0.82, 0.91)** | 0.98 (0.94, 1.02) |
| Diabetes (%) | **1.25 (1.18, 1.32)** | 1.06 (1.00, 1.13) | 0.99 (0.94, 1.05) |
| Heart Failure (%) | **1.08 (1.03, 1.14)** | 1.02 (0.98, 1.06) | **1.13 (1.09, 1.17)** |
| Hypertension (%) | 1.04 (0.95, 1.14) | **1.37 (1.26, 1.49)** | **1.10 (1.02, 1.17)** |
| Kidney Disease (%) | **0.92 (0.87, 0.97)** | 1.02 (0.97, 1.07) | **1.06 (1.02, 1.10)** |
| Stroke (%) | 1.00 (0.95, 1.05) | **1.06 (1.01, 1.11)** | 0.99 (0.95, 1.03) |
| ICU Beds | 0.99 (0.95, 1.04) | 1.04 (1.00, 1.08) | **1.08 (1.05, 1.12)** |
| Nursing Home Beds | **0.96 (0.94, 0.98)** | **1.03 (1.01, 1.05)** | 1.00 (0.98, 1.02) |

**Table S6.** Univariable weekly case fatality rates. Relative risks of county-level variables on weekly case fatality rates (39 repeated measurements per county) by season from 3/23/20-12/21/20. Each row is a separate model and controls for state effects, US census region-specific time varying trends, and additional county overdispersion. Parentheses indicate 95% confidence intervals. Bold indicates confidence interval does not contain 1. Relative risks are for a one standard deviation increase in a variable, except for the metro/nonmetro categorical variable.

| Variable | Univariable Weekly Case Fatality Rate Relative Risk |  |  |
| --- | --- | --- | --- |
|  | Spring | Summer | Fall |
| **Demographic** |  |  |  |
| Age, 20-29 Years (%) | **0.96 (0.94, 0.99)** | **0.86 (0.84, 0.88)** | **0.85 (0.83, 0.86)** |
| Age, 60+ Years (%) | **1.11 (1.08, 1.15)** | **1.15 (1.12, 1.18)** | **1.30 (1.28, 1.33)** |
| Male (%) | **0.67 (0.65, 0.70)** | **0.88 (0.85, 0.91)** | **1.07 (1.04, 1.10)** |
| Metro, >1 million people | Ref | Ref | Ref |
| Metro/Near Metro, <1 million people | 0.94 (0.89, 1.00) | 1.01 (0.96, 1.07) | **1.42 (1.35, 1.49)** |
| Nonmetro, <20,000 people | **0.83 (0.76, 0.90)** | **1.17 (1.09, 1.25)** | **1.91 (1.80, 2.02)** |
| **Race/Ethnicity** |  |  |  |
| Black/African American (%) | **1.14 (1.10, 1.18)** | 0.98 (0.96, 1.01) | **0.81 (0.79, 0.83)** |
| Hispanic/Latino (%) | **0.84 (0.81, 0.86)** | **0.94 (0.92, 0.97)** | **0.79 (0.77, 0.81)** |
| American Indian/Native Alaskan (%) | **0.94 (0.90, 0.98)** | **1.08 (1.04, 1.12)** | **1.06 (1.03, 1.10)** |
| Asian (%) | **0.95 (0.94, 0.97)** | **0.92 (0.90, 0.94)** | **0.80 (0.79, 0.82)** |
| Native Hawaiian/Pacific Islander (%) | **0.90 (0.87, 0.93)** | **0.94 (0.91, 0.96)** | **0.93 (0.90, 0.96)** |
| White/Black Segregation | **1.11 (1.07, 1.15)** | **1.01 (0.98, 1.04)** | **0.86 (0.84, 0.88)** |
| White/non-White Segregation | **1.12 (1.09, 1.16)** | 1.03 (1.00, 1.07) | **0.91 (0.89, 0.94)** |
| **Socioeconomic** |  |  |  |
| Average Household Size | **0.89 (0.87, 0.91)** | 0.98 (0.96, 1.00) | **0.85 (0.84, 0.87)** |
| No Health Insurance (%) | 0.99 (0.95, 1.03) | **1.18 (1.14, 1.23)** | **1.07 (1.03, 1.11)** |
| Poverty (%) | **1.10 (1.07, 1.13)** | **1.11 (1.08, 1.14)** | **1.10 (1.07, 1.12)** |
| No High School Diploma (%) | 1.01 (0.98, 1.04) | **1.16 (1.13, 1.18)** | **1.05 (1.03, 1.08)** |
| Education, Healthcare, Social Worker (%) | 1.00 (0.98, 1.03) | 0.98 (0.96, 1.01) | **1.04 (1.02, 1.06)** |
| **Health** |  |  |  |
| Smokers (%) | **1.07 (1.04, 1.10)** | **1.05 (1.01, 1.08)** | **1.18 (1.15, 1.21)** |
| Obesity (%) | 0.98 (0.96, 1.01) | **1.05 (1.03, 1.08)** | **1.19 (1.16, 1.21)** |
| Asthma (%) | **1.02 (0.99, 1.05)** | 1.00 (0.98, 1.03) | **0.89 (0.87, 0.91)** |
| Cancer (%) | **1.04 (1.01, 1.07)** | 0.98 (0.95, 1.00) | **0.89 (0.87, 0.92)** |
| COPD (%) | **1.05 (1.01, 1.08)** | **1.10 (1.07, 1.13)** | **1.21 (1.18, 1.24)** |
| Diabetes (%) | **1.10 (1.07, 1.13)** | **1.20 (1.17, 1.23)** | **1.01 (0.99, 1.04)** |
| Heart Failure (%) | **1.15 (1.12, 1.19)** | **1.16 (1.13, 1.19)** | **1.09 (1.06, 1.11)** |
| Hypertension (%) | **1.11 (1.07, 1.16)** | **1.33 (1.27, 1.38)** | **1.09 (1.05, 1.14)** |
| Kidney Disease (%) | **1.10 (1.06, 1.13)** | **1.13 (1.10, 1.17)** | 0.98 (0.95, 1.00) |
| Stroke (%) | **1.11 (1.07, 1.15)** | **1.15 (1.11, 1.18)** | **0.92 (0.90, 0.95)** |
| ICU Beds | **1.09 (1.07, 1.12)** | 1.00 (0.98, 1.02) | **0.85 (0.84, 0.87)** |
| Nursing Home Beds | **1.03 (1.01, 1.05)** | 1.02 (1.00, 1.04) | **0.94 (0.93, 0.96)** |

**Table S7.** Multivariable weekly case fatality rates. Relative risks of county-level variables on weekly case fatality rates (39 repeated measurements per county) by season from 3/23/20-12/21/20. All results are from a single model that controls for state effects, US census region-specific time varying trends, and additional county overdispersion. Parentheses indicate 95% confidence intervals. Bold indicates confidence interval does not contain 1. Relative risks are for a one standard deviation increase in a variable, except for the metro/nonmetro categorical variable.

| Variable | Multivariable Weekly Case Fatality Rate Relative Risk |  |  |
| --- | --- | --- | --- |
|  | Spring | Summer | Fall |
| **Demographic** |  |  |  |
| Age, 20-29 Years (%) | 1.00 (0.95, 1.05) | **0.85 (0.82, 0.89)** | **0.96 (0.92, 0.99)** |
| Age, 60+ Years (%) | **1.19 (1.12, 1.27)** | **1.07 (1.02, 1.13)** | **1.10 (1.05, 1.15)** |
| Male (%) | **0.77 (0.73, 0.80)** | **0.93 (0.89, 0.96)** | 0.98 (0.96, 1.01) |
| Metro, >1 million people | Ref | Ref | Ref |
| Metro/Near Metro, <1 million people | 1.01 (0.95, 1.08) | 0.95 (0.89, 1.01) | **1.17 (1.11, 1.23)** |
| Nonmetro, <20,000 people | 0.89 (0.79, 1.01) | 0.94 (0.85, 1.04) | **1.25 (1.16, 1.35)** |
| **Race/Ethnicity** |  |  |  |
| Black/African American (%) | **1.20 (1.13, 1.26)** | 1.00 (0.96, 1.05) | **0.94 (0.90, 0.97)** |
| Hispanic/Latino (%) | **0.79 (0.75, 0.83)** | 0.99 (0.95, 1.04) | **0.95 (0.91, 0.98)** |
| Asian (%) | **1.08 (1.03, 1.14)** | **1.09 (1.04, 1.13)** | 1.01 (0.97, 1.04) |
| American Indian/Native Alaskan (%) | **0.95 (0.92, 0.98)** | 1.02 (0.98, 1.05) | 0.97 (0.95, 1.00) |
| Native Hawaiian/Pacific Islander (%) | 0.97 (0.94, 1.01) | 1.01 (0.98, 1.04) | **1.03 (1.01, 1.06)** |
| White/Black Segregation | 1.00 (0.95, 1.06) | 0.97 (0.93, 1.02) | 0.97 (0.95, 1.00) |
| White/non-White Segregation | **0.94 (0.89, 0.98)** | 1.04 (1.00, 1.08) | 1.02 (0.99, 1.05) |
| **Socioeconomic** |  |  |  |
| Average Household Size | **1.06 (1.02, 1.11)** | **0.93 (0.90, 0.97)** | **0.94 (0.91, 0.97)** |
| No Health Insurance (%) | **0.91 (0.86, 0.97)** | 1.04 (0.98, 1.10) | 1.00 (0.94, 1.05) |
| Poverty (%) | **1.10 (1.04, 1.17)** | 1.04 (0.98, 1.10) | 1.01 (0.97, 1.06) |
| No High School Diploma (%) | 0.98 (0.92, 1.04) | 1.04 (0.99, 1.10) | 1.04 (0.99, 1.09) |
| Education, Healthcare, Social Worker (%) | **0.88 (0.85, 0.91)** | **1.04 (1.01, 1.08)** | 1.03 (1.00, 1.06) |
| **Health** |  |  |  |
| Smokers (%) | 0.97 (0.91, 1.02) | 0.95 (0.90, 1.00) | 0.97 (0.93, 1.02) |
| Obesity (%) | **0.89 (0.86, 0.93)** | 1.00 (0.97, 1.03) | **1.05 (1.02, 1.08)** |
| Asthma (%) | 0.97 (0.94, 1.01) | 0.97 (0.94, 1.00) | 1.00 (0.97, 1.02) |
| Cancer (%) | **0.95 (0.91, 0.99)** | **0.96 (0.92, 0.99)** | 0.99 (0.96, 1.02) |
| COPD (%) | 1.04 (0.98, 1.09) | 0.97 (0.93, 1.01) | 1.02 (0.99, 1.06) |
| Diabetes (%) | **1.09 (1.04, 1.15)** | **1.16 (1.11, 1.22)** | 0.99 (0.94, 1.03) |
| Heart Failure (%) | **1.07 (1.02, 1.12)** | 0.97 (0.93, 1.00) | **1.06 (1.03, 1.10)** |
| Hypertension (%) | **0.87 (0.80, 0.95)** | **1.26 (1.17, 1.35)** | 0.98 (0.92, 1.04) |
| Kidney Disease (%) | **1.06 (1.01, 1.12)** | 0.96 (0.92, 1.00) | **1.07 (1.04, 1.11)** |
| Stroke (%) | 1.00 (0.95, 1.04) | 1.02 (0.98, 1.06) | 1.04 (1.00, 1.07) |
| ICU Beds | **1.13 (1.09, 1.18)** | 1.03 (1.00, 1.07) | 0.99 (0.97, 1.02) |
| Nursing Home Beds | 0.98 (0.97, 1.00) | 1.00 (0.99, 1.02) | 1.00 (0.99, 1.02) |

